# When Two Plus Four Does Not Equal Six: Combining Computational and Functional Evidence to Classify BRCA1 Key Domain Missense Substitutions

**DOI:** 10.1101/2024.10.09.24315186

**Authors:** Scott T. Pew, Madison B. Wiffler, Alun Thomas, Julie L. Boyle, Melissa S. Cline, Nicola J. Camp, David E. Goldgar, Sean V. Tavtigian

## Abstract

Classification of genetic variants remains an obstacle to realizing the full potential of clinical genetic sequencing. Because of their ability to interrogate large numbers of variants, multiplexed assays of variant effect (MAVEs) and computational tools are viewed as a critical part of the solution to variant classification uncertainty. However, the (joint) performance of these assays and tools on novel variants has not been established. Transformation of the qualitative classification guidelines developed by the American College of Medical Genetics and Genomics (ACMG) into a quantitative Bayesian point system enables empirical validation of strength of evidence assigned to evidence criteria. Here, we derived a maximum likelihood estimate (MLE) model that converts frequentist odds ratios calculated from case-control data to proportions pathogenic and applied this model to functional assays, alone and in combination with computational tools across several domains of *BRCA1*. Furthermore, we defined exceptionally conserved ancestral residues (ECARs) and interrogated the performance of assays and tools at these residues in *BRCA1*. We found that missense substitutions in *BRCA1* that fall at ECARs are disproportionately likely to be pathogenic with effect sizes similar to that of protein truncating variants. In contrast, for substitutions falling at non-ECAR positions, concordant predictions of pathogenicity from functional assay and computational tool often fail to meet the additive assumptions of strength in ACMG guidelines. Thus, collectively, we conclude that strengths of evidence assigned by expert opinion in the ACMG guidelines are not universally applicable and require empirical validation.

## INTRODUCTION

ClinVar recently celebrated its 10^th^ anniversary. In that time, as clinical sequencing has become more prevalent, the number of variants cataloged has grown from approximately 27,000 to well over 2 million. Of these 2 million variants, approximately 1.3 million are single nucleotide missense variants (excluding synonymous and nonsense variants), of which roughly 75% remain unclassified (∼990,000 variants) ^1^. Realizing the full potential of clinical genetic sequencing relies on the ability to accurately and efficiently classify these variants into a realistic continuum from pathogenic to benign. To that end, many multiplexed assays of variant effect (MAVE) and computational tools have been developed. However, the accuracy of these assays and tools, used independently and in combination with one another, towards classifying novel variants identified during sequencing remains to be established.

In 2015, the American College of Medical Genetics and Genomics (ACMG) and the Association for Molecular Pathology (AMP) published guidelines that relied on expert opinion to assign strength of evidence to the various evidence criteria used in sequence variant classification ^2^. Subsequently, this approach was found to be compatible with a Bayesian point system where the qualitative assessments of evidence strength categories (supporting, moderate, strong, etc.) were converted to quantitative assertions of odds of pathogenicity (OP) ^3, 4^. Shifting to the Bayesian point system makes possible objective evaluation of the strength of evidence the various evidence criteria actually provide. For example, it was recently demonstrated that several computational tools exceeded the qualitative threshold of “supporting” evidence in favor of benignity or pathogenicity and were able to provide “moderate”, and in some cases “strong” evidence in favor of benignity or pathogenicity ^5^.

In the 2015 ACMG/AMP guidelines, well-established functional assays were qualitatively assigned the ability to provide “strong” evidence in favor of pathogenicity or benignity ^6^. In the Bayesian point system, this translates to +4 points (18.7:1 odds in favor of pathogenicity) or −4 points (0.053:1 odds in favor of pathogenicity). Yet, limited work has been performed to quantitatively evaluate the strength of evidence that functional assays provide. As these assays are not a direct measure of human health, an important step towards applying a specific functional assay’s results to variant classification is validation of the assay’s calibration against human subjects’ data from the disease in question.

Furthermore, the 2015 guidelines provided combining rules that prescribed combinations of different categories of evidence that could move a variant towards classification as pathogenic or benign. Implicit in the original guidelines, and explicitly stated in the subsequent work that transformed the guidelines into a Bayesian framework, is the idea that combining evidence changes the estimate of a sequence variant’s probability of pathogenicity. In a previous publication ^7^, we noted that a majority of variants in the *BRCA1* RING domain could be classified by using computational predictions (ACMG codes: PP3/BP4), whether or not a variant fell at a canonical residue (ACMG code: PM1), functional assay predictions (ACMG codes: PS3/BS3), and minimal human subjects observational data (ACMG codes: PS4, PM2, BS1). This led to our conclusion that validation work is needed to ensure that the strength assigned to these types of evidence was accurate and that addition of Bayesian points associated with these evidence types was the correct mathematical operation.

As a step towards an empirical test, we derive a maximum likelihood estimate (MLE) expression for the proportion of variants that are pathogenic within an analytically defined pool. This model transforms odds ratios (OR) from human subjects’ case-control data into a proportion pathogenic which can then be used to objectively test the strength of evidence a particular category of evidence provides. Subsequently, we demonstrate two important applications for such a model using case-control data from *BRCA1* as a test case. First, we use our model to determine if evidence from several functional assays developed for *BRCA1* generate the strength of evidence assumed in the 2015 ACMG/AMP guidelines. Second, we explore the impact of combining functional assay, canonical residue (which we rename “exceptionally conserved ancestral residues” (ECARs)), and computational tool data to see if doing so changes the proportion pathogenic of analytical subsets of variants in a way that matches the additivity expectations set forth in the Bayesian adaptation of the 2015 classification guidelines (Figure 1). Such an approach allows for the quantitative validation of the qualitative assumptions proposed in variant classification guidelines, a step that is currently lacking for many evidence criteria used in variant classification.

**Figure 1.**
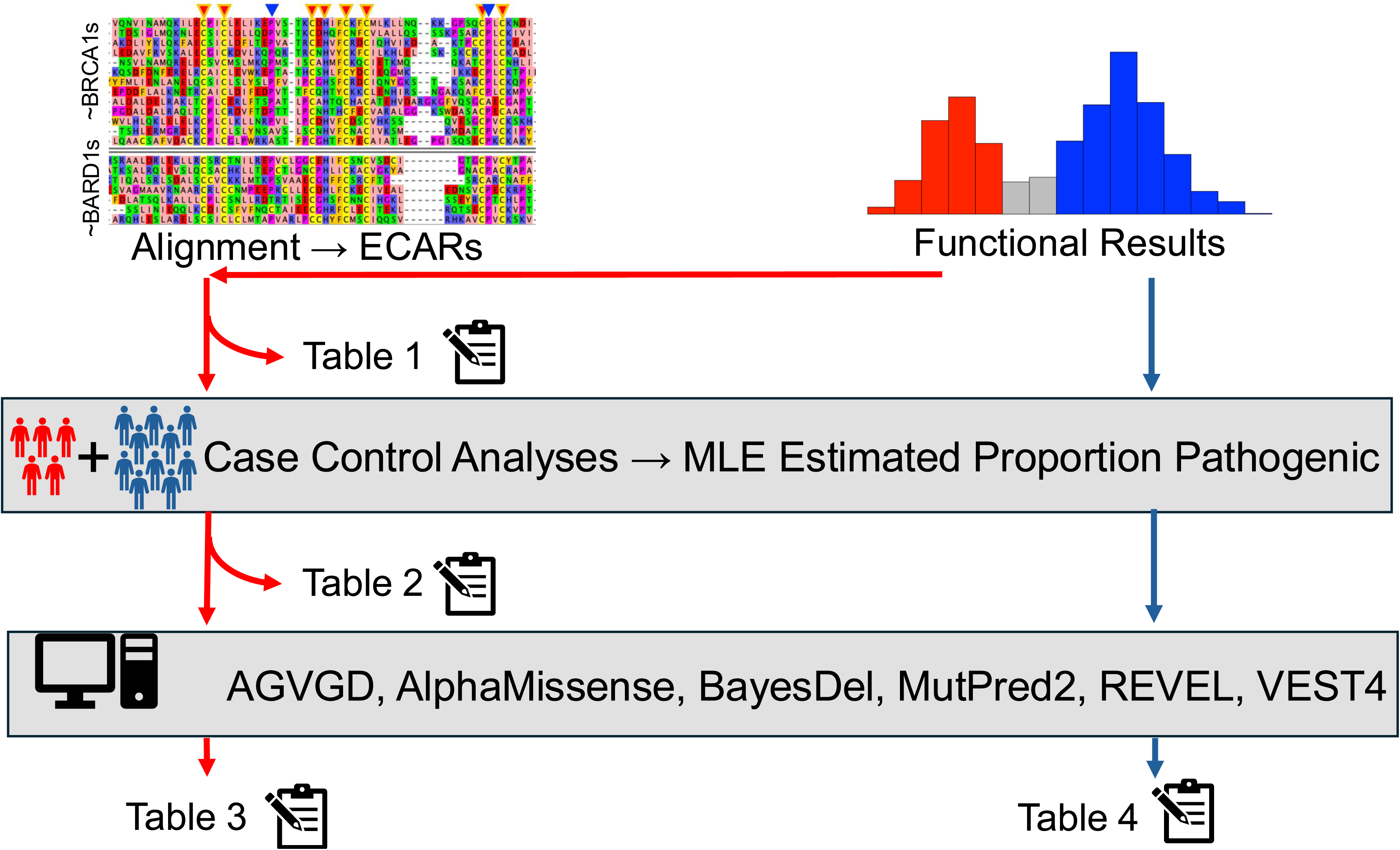
Description of Study Workflow. Functional assay results were combined with other data types (Exceptionally Conserved Ancestral Residue (ECAR) or computational tool prediction) to evaluate the additivity assumptions in the ACMG guidelines. Table 1: Tabulation of the number of functionally abnormal variants (red line) that fall at ECAR positions. Table 2: A case control analysis and conversion to ACMG points is then performed on these variants and results. Table 3: Functionally abnormal variants that do not fall at ECAR positions are combined with predictions from six computational tools, followed by a case control analysis and conversion to ACMG points. Table 4: Functionally normal variants (blue line) from the assay are combined with computational tool predictions from the same six tools, followed by a case control analysis and conversion to ACMG points.

**Table 1.**
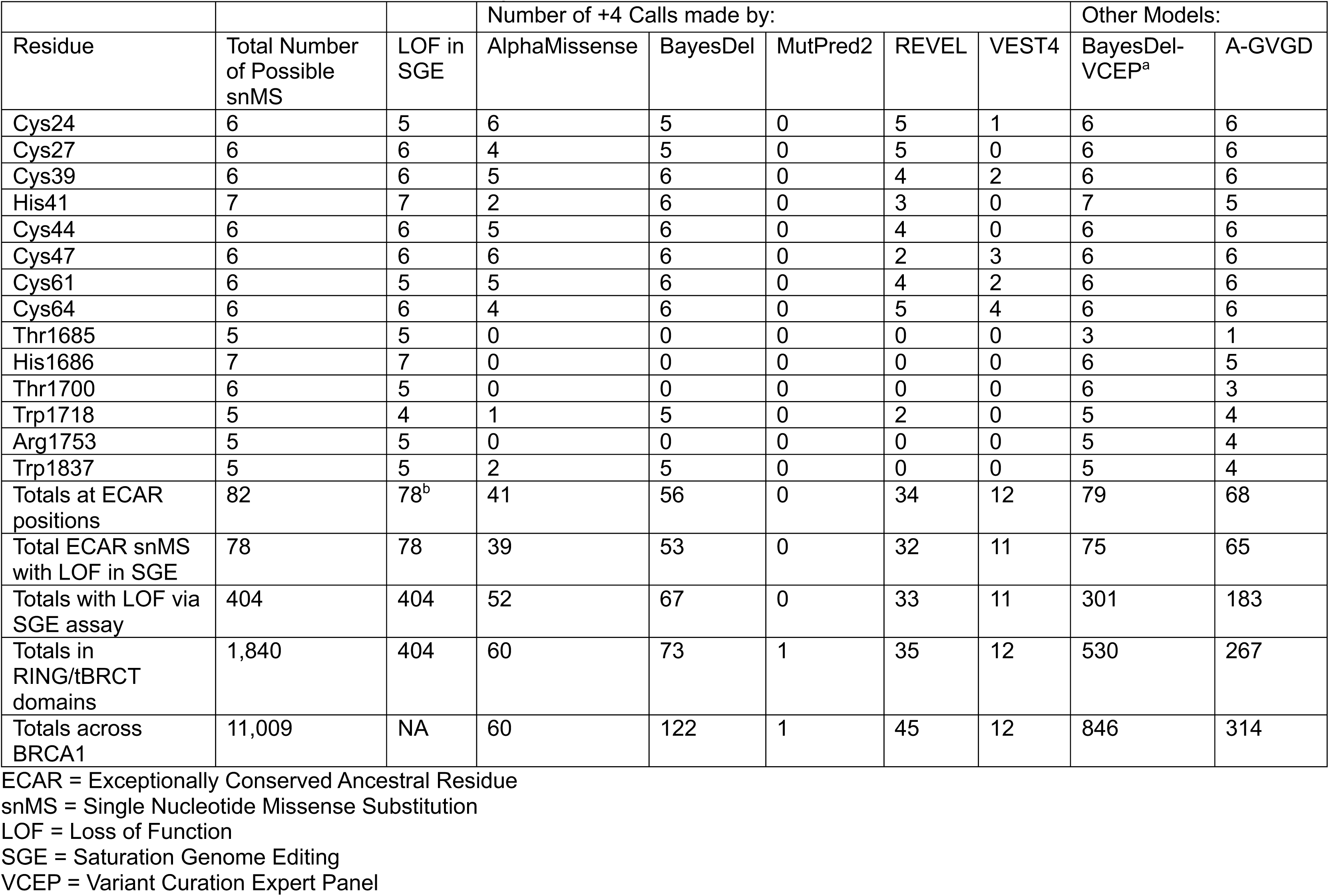
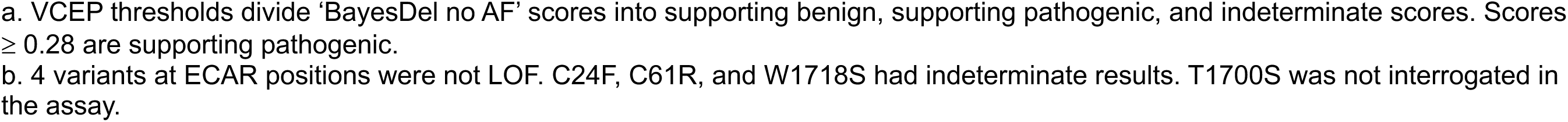
Predictions from SGE Functional Assay and Computational Tools at ECAR Positions.

| Residue | Total Number of Possible snMS | LOF in SGE | Number of +4 Calls made by: |  |  |  |  | Other Models: |  |
| --- | --- | --- | --- | --- | --- | --- | --- | --- | --- |
|  |  |  | AlphaMissense | BayesDel | MutPred2 | REVEL | VEST4 | BayesDel-VCEP<sup>a</sup> | A-GVGD |
| Cys24 | 6 | 5 | 6 | 5 | 0 | 5 | 1 | 6 | 6 |
| Cys27 | 6 | 6 | 4 | 5 | 0 | 5 | 0 | 6 | 6 |
| Cys39 | 6 | 6 | 5 | 6 | 0 | 4 | 2 | 6 | 6 |
| His41 | 7 | 7 | 2 | 6 | 0 | 3 | 0 | 7 | 5 |
| Cys44 | 6 | 6 | 5 | 6 | 0 | 4 | 0 | 6 | 6 |
| Cys47 | 6 | 6 | 6 | 6 | 0 | 2 | 3 | 6 | 6 |
| Cys61 | 6 | 5 | 5 | 6 | 0 | 4 | 2 | 6 | 6 |
| Cys64 | 6 | 6 | 4 | 6 | 0 | 5 | 4 | 6 | 6 |
| Thr1685 | 5 | 5 | 0 | 0 | 0 | 0 | 0 | 3 | 1 |
| His1686 | 7 | 7 | 0 | 0 | 0 | 0 | 0 | 6 | 5 |
| Thr1700 | 6 | 5 | 0 | 0 | 0 | 0 | 0 | 6 | 3 |
| Trp1718 | 5 | 4 | 1 | 5 | 0 | 2 | 0 | 5 | 4 |
| Arg1753 | 5 | 5 | 0 | 0 | 0 | 0 | 0 | 5 | 4 |
| Trp1837 | 5 | 5 | 2 | 5 | 0 | 0 | 0 | 5 | 4 |
| Totals at ECAR positions | 82 | 78<sup>b</sup> | 41 | 56 | 0 | 34 | 12 | 79 | 68 |
| Total ECAR snMS with LOF in SGE | 78 | 78 | 39 | 53 | 0 | 32 | 11 | 75 | 65 |
| Totals with LOF via SGE assay | 404 | 404 | 52 | 67 | 0 | 33 | 11 | 301 | 183 |
| Totals in RING/tBRCT domains | 1,840 | 404 | 60 | 73 | 1 | 35 | 12 | 530 | 267 |
| Totals across BRCA1 | 11,009 | NA | 60 | 122 | 1 | 45 | 12 | 846 | 314 |

ECAR = Exceptionally Conserved Ancestral Residue
snMS = Single Nucleotide Missense Substitution
LOF = Loss of Function
SGE = Saturation Genome Editing
VCEP = Variant Curation Expert Panel
- a. VCEP thresholds divide 'BayesDel no AF' scores into supporting benign, supporting pathogenic, and indeterminate scores. Scores $\geq 0.28$ are supporting pathogenic. - b. 4 variants at ECAR positions were not LOF. C24F, C61R, and W1718S had indeterminate results. T1700S was not interrogated in the assay.

**Table 2.**
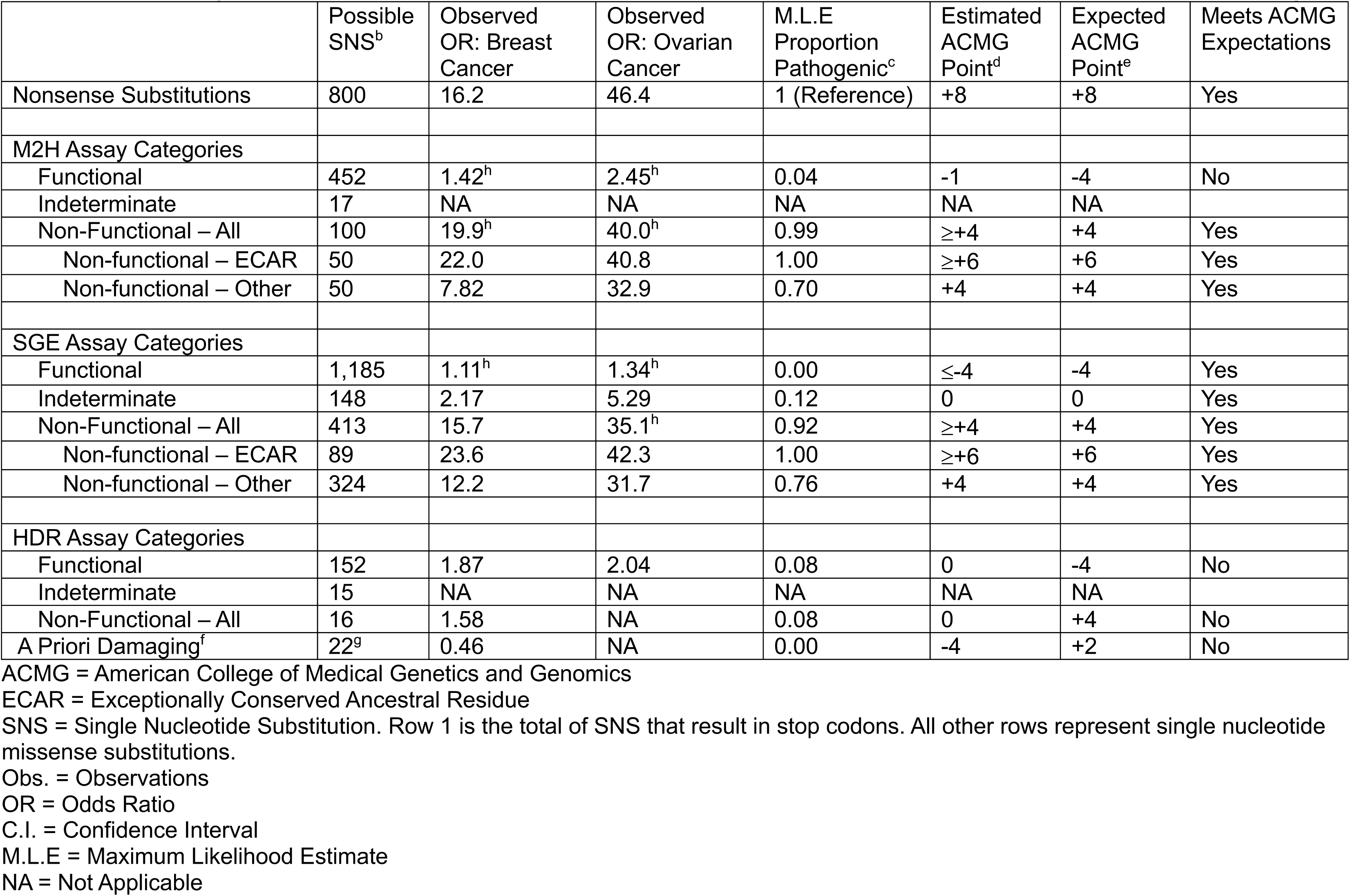
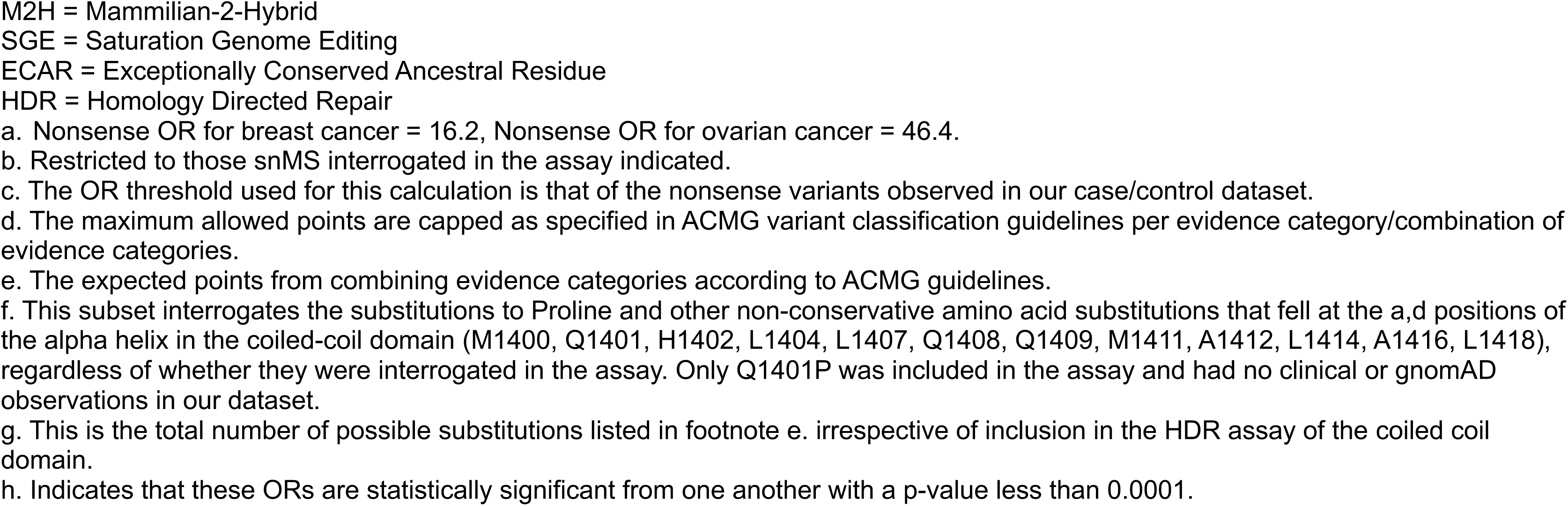
Proportions Pathogenic, and ACMG Point Estimates of Functional Assay Analytical Subsets and Non-functional Substitutions Falling at ECAR Positions. OR Threshold equal to that of nonsense variants in breast and ovarian cancer groups^a^.

|  | Possible SNS<sup>b</sup> | Observed OR: Breast Cancer | Observed OR: Ovarian Cancer | M.L.E Proportion Pathogenic<sup>c</sup> | Estimated ACMG Point<sup>d</sup> | Expected ACMG Point<sup>e</sup> | Meets ACMG Expectations |
| --- | --- | --- | --- | --- | --- | --- | --- |
| Nonsense Substitutions | 800 | 16.2 | 46.4 | 1 (Reference) | +8 | +8 | Yes |
| M2H Assay Categories |  |  |  |  |  |  |  |
| Functional | 452 | 1.42<sup>h</sup> | 2.45<sup>h</sup> | 0.04 | -1 | -4 | No |
| Indeterminate | 17 | NA | NA | NA | NA | NA |  |
| Non-Functional – All | 100 | 19.9<sup>h</sup> | 40.0<sup>h</sup> | 0.99 | ≥+4 | +4 | Yes |
| Non-functional – ECAR | 50 | 22.0 | 40.8 | 1.00 | ≥+6 | +6 | Yes |
| Non-functional – Other | 50 | 7.82 | 32.9 | 0.70 | +4 | +4 | Yes |
| SGE Assay Categories |  |  |  |  |  |  |  |
| Functional | 1,185 | 1.11<sup>h</sup> | 1.34<sup>h</sup> | 0.00 | ≤-4 | -4 | Yes |
| Indeterminate | 148 | 2.17 | 5.29 | 0.12 | 0 | 0 | Yes |
| Non-Functional – All | 413 | 15.7 | 35.1<sup>h</sup> | 0.92 | ≥+4 | +4 | Yes |
| Non-functional – ECAR | 89 | 23.6 | 42.3 | 1.00 | ≥+6 | +6 | Yes |
| Non-functional – Other | 324 | 12.2 | 31.7 | 0.76 | +4 | +4 | Yes |
| HDR Assay Categories |  |  |  |  |  |  |  |
| Functional | 152 | 1.87 | 2.04 | 0.08 | 0 | -4 | No |
| Indeterminate | 15 | NA | NA | NA | NA | NA |  |
| Non-Functional – All | 16 | 1.58 | NA | 0.08 | 0 | +4 | No |
| A Priori Damaging<sup>f</sup> | 22<sup>g</sup> | 0.46 | NA | 0.00 | -4 | +2 | No |

ACMG = American College of Medical Genetics and Genomics
ECAR = Exceptionally Conserved Ancestral Residue
SNS = Single Nucleotide Substitution. Row 1 is the total of SNS that result in stop codons. All other rows represent single nucleotide missense substitutions.
Obs. = Observations
OR = Odds Ratio
C.I. = Confidence Interval
M.L.E = Maximum Likelihood Estimate
NA = Not Applicable
M2H = Mammalian-2-Hybrid
SGE = Saturation Genome Editing
HDR = Homology Directed Repair
a. Nonsense OR for breast cancer = 16.2, Nonsense OR for ovarian cancer = 46.4.

**Table 3.**
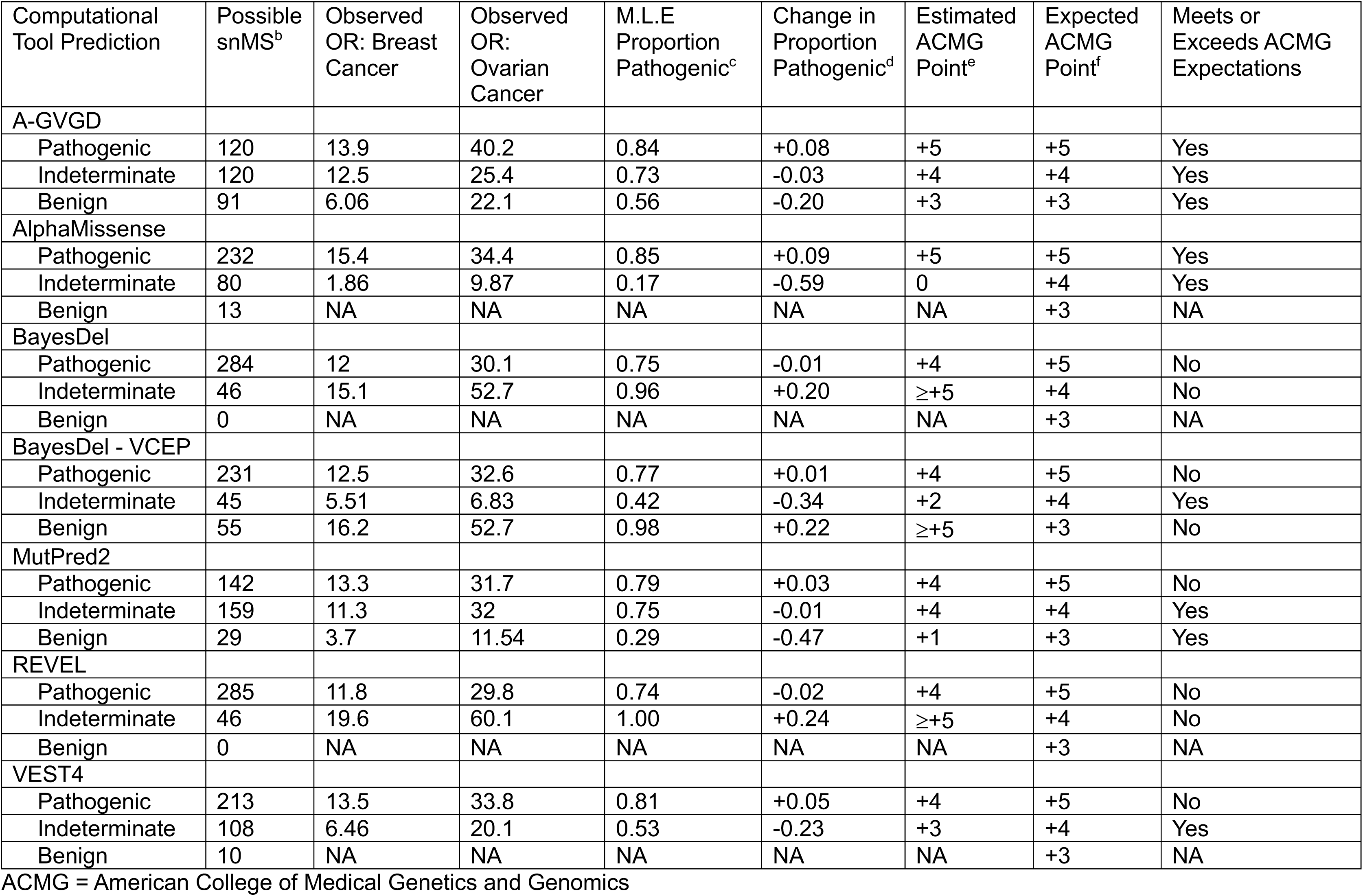
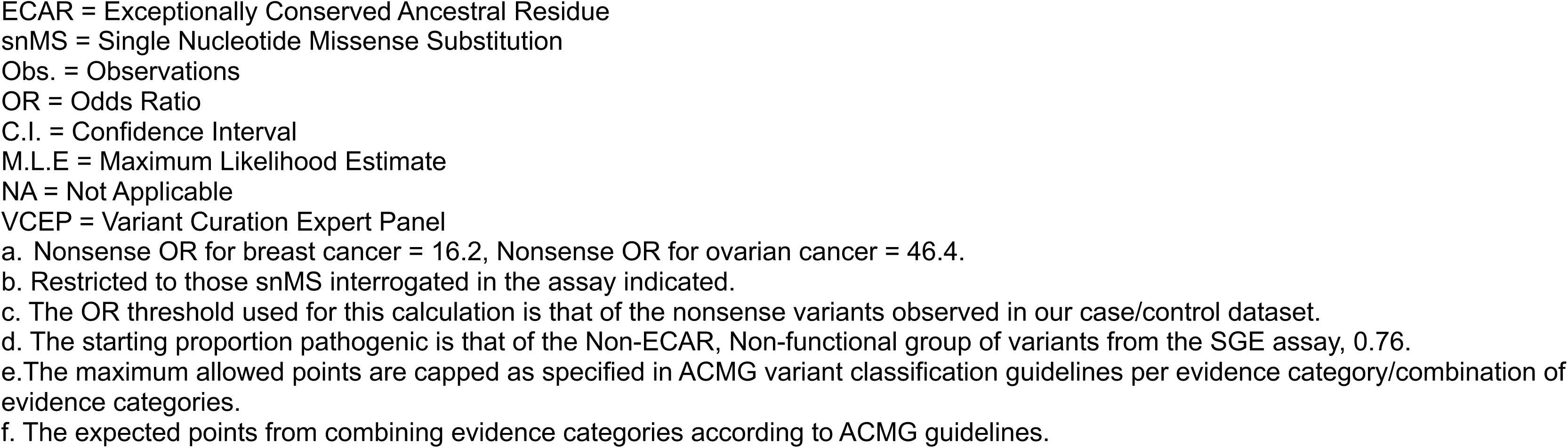
Proportions Pathogenic, and ACMG Point Estimates of Non-functional, Non-ECAR Assay Results Combined with Computational Predictions. OR Threshold equal to that of nonsense variants in breast and ovarian cancer groups^a^.

| Computational Tool Prediction | Possible snMS<sup>b</sup> | Observed OR: Breast Cancer | Observed OR: Ovarian Cancer | M.L.E Proportion Pathogenic<sup>c</sup> | Change in Proportion Pathogenic<sup>d</sup> | Estimated ACMG Point<sup>e</sup> | Expected ACMG Point<sup>f</sup> | Meets or Exceeds ACMG Expectations |
| --- | --- | --- | --- | --- | --- | --- | --- | --- |
| A-GVGD |  |  |  |  |  |  |  |  |
| Pathogenic | 120 | 13.9 | 40.2 | 0.84 | +0.08 | +5 | +5 | Yes |
| Indeterminate | 120 | 12.5 | 25.4 | 0.73 | -0.03 | +4 | +4 | Yes |
| Benign | 91 | 6.06 | 22.1 | 0.56 | -0.20 | +3 | +3 | Yes |
| AlphaMissense |  |  |  |  |  |  |  |  |
| Pathogenic | 232 | 15.4 | 34.4 | 0.85 | +0.09 | +5 | +5 | Yes |
| Indeterminate | 80 | 1.86 | 9.87 | 0.17 | -0.59 | 0 | +4 | Yes |
| Benign | 13 | NA | NA | NA | NA | NA | +3 | NA |
| BayesDel |  |  |  |  |  |  |  |  |
| Pathogenic | 284 | 12 | 30.1 | 0.75 | -0.01 | +4 | +5 | No |
| Indeterminate | 46 | 15.1 | 52.7 | 0.96 | +0.20 | ≥+5 | +4 | No |
| Benign | 0 | NA | NA | NA | NA | NA | +3 | NA |
| BayesDel - VCEP |  |  |  |  |  |  |  |  |
| Pathogenic | 231 | 12.5 | 32.6 | 0.77 | +0.01 | +4 | +5 | No |
| Indeterminate | 45 | 5.51 | 6.83 | 0.42 | -0.34 | +2 | +4 | Yes |
| Benign | 55 | 16.2 | 52.7 | 0.98 | +0.22 | ≥+5 | +3 | No |
| MutPred2 |  |  |  |  |  |  |  |  |
| Pathogenic | 142 | 13.3 | 31.7 | 0.79 | +0.03 | +4 | +5 | No |
| Indeterminate | 159 | 11.3 | 32 | 0.75 | -0.01 | +4 | +4 | Yes |
| Benign | 29 | 3.7 | 11.54 | 0.29 | -0.47 | +1 | +3 | Yes |
| REVEL |  |  |  |  |  |  |  |  |
| Pathogenic | 285 | 11.8 | 29.8 | 0.74 | -0.02 | +4 | +5 | No |
| Indeterminate | 46 | 19.6 | 60.1 | 1.00 | +0.24 | ≥+5 | +4 | No |
| Benign | 0 | NA | NA | NA | NA | NA | +3 | NA |
| VEST4 |  |  |  |  |  |  |  |  |
| Pathogenic | 213 | 13.5 | 33.8 | 0.81 | +0.05 | +4 | +5 | No |
| Indeterminate | 108 | 6.46 | 20.1 | 0.53 | -0.23 | +3 | +4 | Yes |
| Benign | 10 | NA | NA | NA | NA | NA | +3 | NA |

ACMG = American College of Medical Genetics and Genomics
ECAR = Exceptionally Conserved Ancestral Residue
snMS = Single Nucleotide Missense Substitution
Obs. = Observations
OR = Odds Ratio
C.I. = Confidence Interval
M.L.E = Maximum Likelihood Estimate
NA = Not Applicable
VCEP = Variant Curation Expert Panel
a. Nonsense OR for breast cancer = 16.2, Nonsense OR for ovarian cancer = 46.4.
b. Restricted to those snMS interrogated in the assay indicated.

**Table 4.**
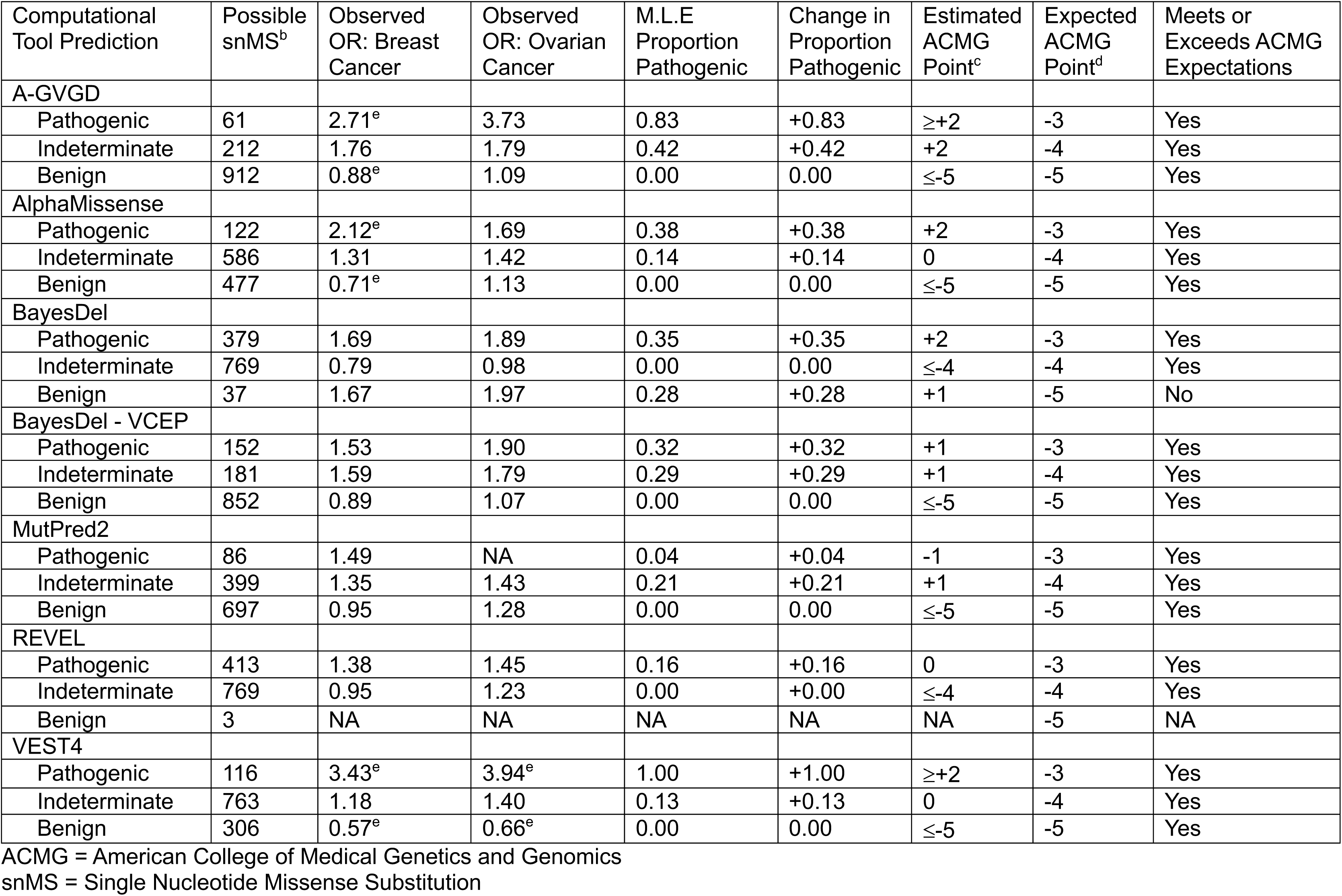
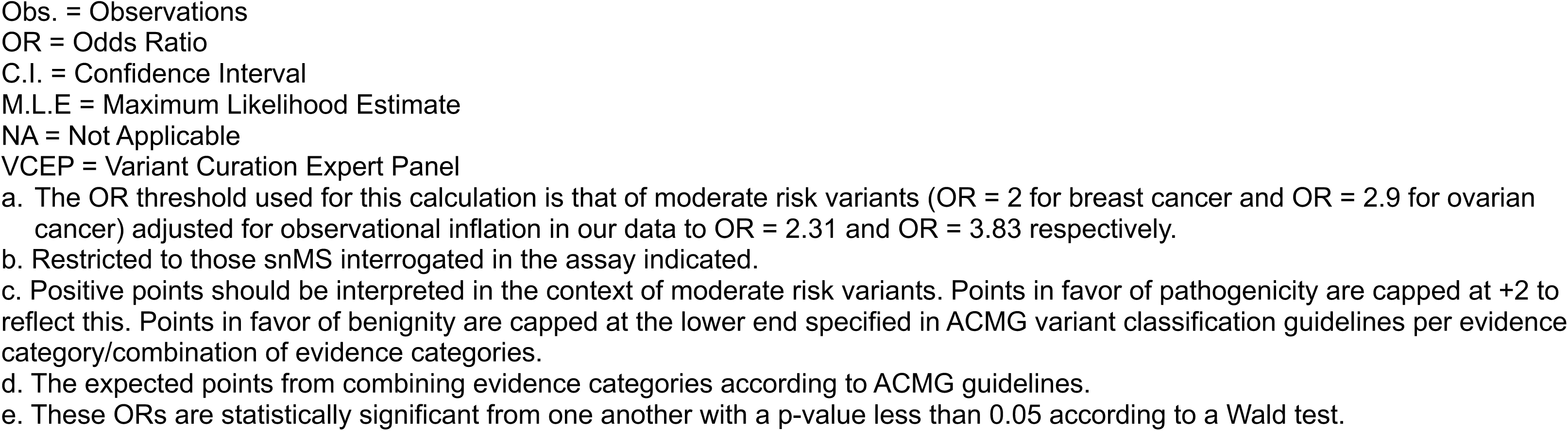
Proportions Pathogenic, and ACMG Point Estimates of Functionally Normal Assay Results Combined with Computational Predictions. OR threshold equal to that of moderate risk variants^a^.

| Computational Tool Prediction | Possible snMS<sup>b</sup> | Observed OR: Breast Cancer | Observed OR: Ovarian Cancer | M.L.E Proportion Pathogenic | Change in Proportion Pathogenic | Estimated ACMG Point<sup>c</sup> | Expected ACMG Point<sup>d</sup> | Meets or Exceeds ACMG Expectations |
| --- | --- | --- | --- | --- | --- | --- | --- | --- |
| A-GVGD |  |  |  |  |  |  |  |  |
| Pathogenic | 61 | 2.71<sup>e</sup> | 3.73 | 0.83 | +0.83 | ≥+2 | -3 | Yes |
| Indeterminate | 212 | 1.76 | 1.79 | 0.42 | +0.42 | +2 | -4 | Yes |
| Benign | 912 | 0.88<sup>e</sup> | 1.09 | 0.00 | 0.00 | ≤-5 | -5 | Yes |
| AlphaMissense |  |  |  |  |  |  |  |  |
| Pathogenic | 122 | 2.12<sup>e</sup> | 1.69 | 0.38 | +0.38 | +2 | -3 | Yes |
| Indeterminate | 586 | 1.31 | 1.42 | 0.14 | +0.14 | 0 | -4 | Yes |
| Benign | 477 | 0.71<sup>e</sup> | 1.13 | 0.00 | 0.00 | ≤-5 | -5 | Yes |
| BayesDel |  |  |  |  |  |  |  |  |
| Pathogenic | 379 | 1.69 | 1.89 | 0.35 | +0.35 | +2 | -3 | Yes |
| Indeterminate | 769 | 0.79 | 0.98 | 0.00 | 0.00 | ≤-4 | -4 | Yes |
| Benign | 37 | 1.67 | 1.97 | 0.28 | +0.28 | +1 | -5 | No |
| BayesDel - VCEP |  |  |  |  |  |  |  |  |
| Pathogenic | 152 | 1.53 | 1.90 | 0.32 | +0.32 | +1 | -3 | Yes |
| Indeterminate | 181 | 1.59 | 1.79 | 0.29 | +0.29 | +1 | -4 | Yes |
| Benign | 852 | 0.89 | 1.07 | 0.00 | 0.00 | ≤-5 | -5 | Yes |
| MutPred2 |  |  |  |  |  |  |  |  |
| Pathogenic | 86 | 1.49 | NA | 0.04 | +0.04 | -1 | -3 | Yes |
| Indeterminate | 399 | 1.35 | 1.43 | 0.21 | +0.21 | +1 | -4 | Yes |
| Benign | 697 | 0.95 | 1.28 | 0.00 | 0.00 | ≤-5 | -5 | Yes |
| REVEL |  |  |  |  |  |  |  |  |
| Pathogenic | 413 | 1.38 | 1.45 | 0.16 | +0.16 | 0 | -3 | Yes |
| Indeterminate | 769 | 0.95 | 1.23 | 0.00 | +0.00 | ≤-4 | -4 | Yes |
| Benign | 3 | NA | NA | NA | NA | NA | -5 | NA |
| VEST4 |  |  |  |  |  |  |  |  |
| Pathogenic | 116 | 3.43<sup>e</sup> | 3.94<sup>e</sup> | 1.00 | +1.00 | ≥+2 | -3 | Yes |
| Indeterminate | 763 | 1.18 | 1.40 | 0.13 | +0.13 | 0 | -4 | Yes |
| Benign | 306 | 0.57<sup>e</sup> | 0.66<sup>e</sup> | 0.00 | 0.00 | ≤-5 | -5 | Yes |

ACMG = American College of Medical Genetics and Genomics
snMS = Single Nucleotide Missense Substitution
Obs. = Observations
OR = Odds Ratio
C.I. = Confidence Interval
M.L.E = Maximum Likelihood Estimate
NA = Not Applicable
VCEP = Variant Curation Expert Panel
- a. The OR threshold used for this calculation is that of moderate risk variants (OR = 2 for breast cancer and OR = 2.9 for ovarian cancer) adjusted for observational inflation in our data to OR = 2.31 and OR = 3.83 respectively. - b. Restricted to those snMS interrogated in the assay indicated. - c. Positive points should be interpreted in the context of moderate risk variants. Points in favor of pathogenicity are capped at +2 to reflect this. Points in favor of benignity are capped at the lower end specified in ACMG variant classification guidelines per evidence category/combination of evidence categories. - d. The expected points from combining evidence categories according to ACMG guidelines. - e. These ORs are statistically significant from one another with a p-value less than 0.05 according to a Wald test.

## METHODS

### Maximum Likelihood Estimate of Proportion Pathogenic

In addition to the assumptions in the Bayesian point system (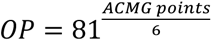, prior probability of pathogenicity = 0.102, points are log odds and therefore additive), this approach relies on several other assumptions:

1. The OR of a pool of truncating variants in loss of function susceptibility genes provides a “standard candle” by which one can measure the magnitude of effect of a given analytically defined pool of variants.
2. An analytically defined pool of variants with an OR similar to that of a pool of truncating variants consists almost entirely of variants that are pathogenic with an effect size approximately equal to that of the truncating variants.
3. An analytically defined pool of variants with an OR near 1.0 consists almost entirely of benign variants.
4. Analytically defined pools of variants with an OR between 1.0 and the OR of truncating variants can be modeled as a two-component mixture of benign and pathogenic variants. This assumption is debatable, but importantly, is also testable.

The full derivation of the maximum likelihood estimate model is presented in the Supplemental Methods. In brief, our model estimates the proportion of observed variants that are pathogenic, *q*, from the proportion of all variants that are pathogenic, *p*, the number of cases, *n*, the number of controls, *m*, the population frequency of disease among individuals who do not carry a pathogenic variant, *δ*, and the frequency of disease among individuals who are carriers of a pathogenic variant, *δ_1_*, as shown in Equation 1:

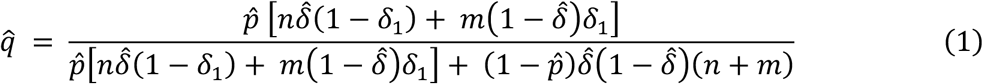

In addition to the variables listed above, our MLE model requires input values for an odds ratio threshold at which all variants are assumed to be pathogenic. For the odds ratio threshold in our analysis, truncating variants observed in our case-control dataset were tabulated and frequentist odds ratios for breast and ovarian cancers were calculated for these groups of variants (16.2, 95% CI: 12.7 – 20.8 for breast cancer and 46.4, 95% CI: 35.7 – 60.7 for ovarian cancer). To evaluate the prediction of benign variants we used an alternative OR ceiling chosen to match the threshold of moderate-risk variants (OR = 2.0 for breast cancer and OR = 2.9 for ovarian cancer), which was calculated to be inflated to 2.31 and 3.83 in our family history enriched case-control dataset ^8, 9^. The general population frequency of cancer used in the analysis was 0.125 for breast cancer, and 0.011 for ovarian cancer ^10, 11^.

### Identification of Exceptionally Conserved Ancestral Residues in BRCA1 / BARD1

To find evolutionarily distant BRCA1/BARD1 homologs, we started with two alignments, one of the RING domains of BRCA1 and BARD1 from *Homo sapiens* and *Strongylocentrotus purpuratus* and a second of the corresponding tandem BRCT (tBRCT) domains. These alignments were prepared using the structure-guided 3D-Coffee/Expresso mode from the T-coffee suite of sequence alignment tools ^12, 13^. The two resulting alignments were used with hmmsearch to scan phylogenetically defined subsets of the reference proteome database maintained at the HMMER website ^14^. Hits were defined as gene models with high-scoring matches to both the RING and tandem BRCT alignments.

At the first round, hits from Placozoa and single celled Holozoans were included, the alignments rebuilt, and then trimmed back to include clear *BRCA1* and *BARD1* orthologs from *Trichoplax* and *Salpingoeca*. The process was iterated to obtain and include homologs from the Fungi and then reiterated to obtain and include homologs from three additional eukaryotic supergroups: Archaeplastida, SAR, and Discoba.

To prepare the final alignments and then define exceptionally conserved ancestral residues (ECARs), we stratified the hits into a group of *BARD1* and relatively *BARD1*-like hits and a second group of *BRCA1* and relatively *BRCA1*-like groups; ambiguous hits were placed in the *BRCA1*-like group. As before, RING and tBRCT alignments were prepared using 3D-Coffee/Expresso. Then the BRCA1 and BARD1 alignments were aligned to each other by introducing gaps across the whole BRCA1 or whole BARD1 alignment, i.e., without modifying the within-group alignments. The tBRCT alignments were then appended to the RING domain alignments. Finally, ECARs were defined as (a) positions that were invariant in our existing metazoan BRCA1 and BARD1 alignments used with Align-GVGD ^15, 16^, and either (b) also invariant in the new multi-supergroup alignments or (c) had just one conservative substitution (i.e., Grantham Variation <65) in the new multi-supergoup alignment.

### Functional Assay Results and Computational Predictions

Functional assay scores were obtained from three separate assays that interrogate missense variants in various domains of *BRCA1*. These assays were: the 2018 Findlay et al. saturation genome editing (SGE) assay that interrogated missense and splice junction variants in the RING and tBRCT domain of *BRCA1* ^17^, the 2022 Clark et al. mammalian-2-hybrid (M2H) assay performed by our laboratory that evaluated missense variants in the RING domain of *BRCA1* ^7^, and the 2023 Nagy et al. homology-directed repair (HDR) assay that evaluated missense variants in the coiled-coil domain of *BRCA1* ^18^. The M2H and SGE assays were selected due to their nearly comprehensive evaluation of all possible snMS variants in the RING and/or tBRCT domains of BRCA1. The HDR assay was selected because it interrogated the coiled-coil domain which is considered a clinically import functional domain^19^. Scores for these assays were downloaded from MaveDB (SGE and M2H) ^20^ or from the publication ^18^. Thresholds for functional, indeterminate and non-functional variants were used as defined by the relevant publication. We used the calibrated scores for each assay in our analysis because these are the thresholds that are likely to be used during clinical variant classification.

Computational tool scores for BayesDel no-AF ^21^, REVEL ^22^, and VEST4 ^23^ were obtained from the database for Non-synonymous Functional Predictions (dbNSFP) via Ensembl’s web-based Variant Effect Predictor (VEP) ^24, 25^. Computational scores for MutPred2 were obtained by downloading a local copy of the software^26^. Scores for AlphaMissense were downloaded from the original publication ^27^. Scores for Align-GVGD (A-GVGD) ^28, 29^ were obtained from agvgd.hci.utah.edu using the Human to Sea Urchin alignment. Thresholds for pathogenic and benign for AlphaMissense, BayesDel, MutPred2, REVEL, and VEST4 were the supporting pathogenic and supporting benign thresholds from recent calibrations of these tools^5, 30^. Align-GVGD, predictions with a score of C0 were interpreted as predicted benign, intermediate scores as indeterminate, and scores of C65 were interpreted as predicted pathogenic. AlphaMissense, BayesDel, MutPred2, REVEL, and VEST4 were selected because they demonstrated the ability to provide strong evidence in favor of pathogenicity in recent calibration studies^5, 30^ and some of these tools are often used by Variant Curation Expert Panels (VCEP). While the scales for each tool may differ, the individual thresholds used in this work are those established in Pejaver et al. to be sufficient for supporting evidence for both benign and pathogenic variants in the ACMG classification framework. A-GVGD has previously been demonstrated to perform well in *BRCA1*^15^.

### Estimate of ACMG Points

We interpreted the MLE proportion pathogenic as a posterior probability, PP (represented as *q* in the supplemental text and Equation 1), of the variants in the analytical subset being pathogenic and we thus converted this estimate to the appropriate ACMG point value via an OP using a prior probability (P1) of 0.102^4^. Proportions pathogenic were calculated separately for each cancer type using our MLE model, converted to odds of pathogenicity using Equation 2 below, and then combined using a weighted geometric mean based on the number of individuals in the breast cancer case control group and ovarian cancer case control group.

This new estimate of odds of pathogenicity was subsequently converted back to a proportion pathogenic to display in Tables 2 – 4 and converted to ACMG points using Equation 3^4^. When assigning ACMG points from the estimated proportion pathogenic, the points that could be awarded were limited to the maximum values prescribed in the ACMG guidelines unless otherwise noted ^2, 4^.

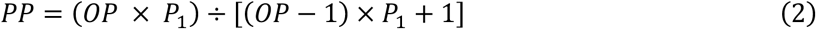

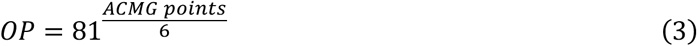

### Case and Control Count Sources and Odds Ratio Calculations

Human subjects’ data were obtained from Ambry Genetics and Myriad Genetics. Control counts are from the non-cancer cohort of gnomAD v2 exomes and gnomAD v3 genomes^31, 32^. Ethnicity for this dataset was split into four groups: non-Finnish European (NFE), Ashkenazi Jewish, Finnish, and all others. Ashkenazi Jewish and Finnish individuals were removed due to strong founder effects in these populations. Individuals from Ambry and Myriad were considered breast cancer cases if they had breast cancer and did not have ovarian cancer. Individuals from Ambry and Myriad were considered ovarian cancer cases if they either had ovarian cancer or ovarian and breast cancer.

The Ambry dataset initially included data generated by Ambry Genetics from multi-gene sequencing panels for 165,031 individuals “exempted from review by the Western Institutional Review Board” ^33^. Filters were applied to remove those individuals without breast or ovarian cancer, individuals who lacked *BRCA1* sequence data, individuals of Ashkenazi Jewish or Finnish ancestry, and individuals with pathogenic or likely pathogenic variants in *TP53*, *BRCA2*, and *PALB2*. This resulted in a final dataset of 98,111 individuals, 66,672 (68%) of which were of NFE ethnicity and 31,439 (32%) of other ethnicity. Of individuals of NFE ethnicity, 57,431 (86%) had breast cancer and 9,241 (14%) had ovarian cancer. Of individuals of other ethnicity, 28,114 (89%) had breast cancer and 3,325 (11%) had ovarian cancer. 2,970 individuals had variants in *BRCA1*: 1,266 (43%) individuals of NFE ethnicity who had breast cancer, 452 (15%) individuals of NFE ethnicity who had ovarian cancer, 1,013 (34%) individuals of other ethnicity who had breast cancer, and 239 (8%) individuals of other ethnicity who had ovarian cancer. 477 individuals had missense variants that fell within the RING (amino acids 1-100), coiled-coil (amino acids 1,280-1,576), or tBRCT domain (amino acids 1,650-1,863): 118 (25%) individuals of NFE ethnicity who had breast cancer, 155 (32%) individuals of other ethnicity who had breast cancer, 123 (26%) individuals of NFE ethnicity who had ovarian cancer, and 81(17%) individuals of other ethnicity who had ovarian cancer.

The Myriad dataset initially included data generated by full-sequence BRCA1/2 tests performed by Myriad Genetics from 60,607 individuals. For these, a test request form must have been completed by the ordering health care provider, and the form must have been signed by an appropriate individual indicating that “informed consent has been signed and is on file” ^15, 33, 34^. Filters were applied to remove those individuals without breast or ovarian cancer, individuals who lacked *BRCA1* sequence data, individuals of Ashkenazi Jewish or Finnish ancestry, and individuals with pathogenic or likely pathogenic variants in *BRCA2*. This resulted in a final dataset of 35,088 individuals, 25,653 (73%) of which were of NFE ethnicity and 9,435 (27%) of other ethnicity. Of individuals of NFE ethnicity, 22,504 (88%) had breast cancer and 3,149 (12%) had ovarian cancer. Of individuals of other ethnicity, 8,347 (88%) had breast cancer and 1,088 (12%) had ovarian cancer. 4,005 individuals had variants in *BRCA1*: 2,231(56%) individuals of NFE ethnicity who had breast cancer, 639 (16%) individuals of NFE ethnicity who had ovarian cancer, 904 (22%) individuals of other ethnicity who had breast cancer, and 231 (6%) individuals of other ethnicity who had ovarian cancer. 524 had missense variants that fell within the RING, coiled-coil, or tBRCT domain: 289 (55%) individuals of NFE ethnicity who had breast cancer, 83 (16%) individuals of NFE ethnicity who had ovarian cancer, 124 (24%) individuals of other ethnicity who had ovarian cancer, and 28 (5%) individuals of other ethnicity who had ovarian cancer.

The gnomAD dataset is a composite of non-overlapping v2 non-cancer exomes and v3 non-cancer genomes that initially included 192,502 individuals. Filters were applied to remove individuals of Finnish or Ashkenazi Jewish ancestry and to remove common missense variants (allele frequency > 0.000271). This resulted in a final dataset of 169,933 individuals: 84,396 (50%) individuals of NFE ethnicity and 85,537 (50%) individuals of other ethnicity. 644 individuals had missense variants that fell within the *BRCA1* RING, coiled-coil, or tBRCT domain: 264 (41%) individuals of NFE ethnicity and 380 (59%) individuals of other ethnicity.

Combining these datasets resulted in a final dataset of 133,199 breast and ovarian cases from Ambry and Myriad and 169,933 controls from gnomAD v2 and v3.

After filtering, the data for *BRCA1* were formatted into a table that indicated the ethnicity of the individual, whether an individual was a case or control, how the variant was classified by each functional assay and computational tool, and whether the variant fell at an ECAR.

Logistic regression was performed using R studio to calculate frequentist odds ratios for each stratum (functional, indeterminate, loss-of-function (LOF) all, LOF-ECAR, LOF-other) of the functional assays included in our analysis. Odds ratios were calculated similarly for strata that resulted in combining SGE functional assay results and computational tool predictions. Proportions pathogenic were calculated using a custom R script. Following logistic regression, the Wald test was performed to determine statistical significance between strata in each analysis.

To determine if combining computational tool predictions to functional assay predictions was statistically significant, multivariate logistic regression was performed using combinations of the SGE functional assay result (functional or LOF) and computational tool predictions (benign or pathogenic) for each tool analyzed in our study.

## RESULTS

### Exceptionally Conserved Ancestral Residues in *BRCA1* / *BARD1*

*BRCA1*, *BARD1*, and homologs that have been studied in the Archaeplastida are thought to have evolved from a common ancestor gene with a 5’ RING domain and 3’ tandem BRCT (tBRCT) domain that was present in an ancient eukaryote before the divergence of Amorphea and Archaeplastida ^35^. Protein multiple sequence alignment (PMSA) based hmmsearch ^14^ identified clear *BRCA1* and *BARD1* orthologs in *Trichoplax* and more distantly related single-celled members of Holozoa, plus homologs (some of which may be true orthologs) in the Holomycota, Archaeplastida, SAR, and Discoba. Although the root of the eukaryotic phylogeny remains unclear ^36, 37^, the resulting PMSA should be sufficient to identify positions in human *BRCA1* and *BARD1* where the amino acid present in the two human sequences has been inherited identical by descent from a common ancestor gene and is under constraint in the Amorphea and 2-3 additional eukaryotic supergroups. We identified 14 such residues: the 8 residues that define the C3HC4 RING motif plus 6 additional residues in the tBRCT domain (Table 1, Figure S1), and hereafter refer to them as Exceptionally Conserved Ancestral Residues (ECARs).

Of the 82 possible snMSs at ECARs, 78 resulted in a non-functional assay result via the SGE assay. The four exceptions were C24F, C61R, T1700S (which was not interrogated in the SGE assay), and W1718S. C24F, C61R, and W1718S had indeterminate results in the SGE assay, however, C24F and C61R were scored as loss of function in the M2H assay.

Using the pathogenic strong thresholds established in Pejaver et al, we also tabulated the number of substitutions at ECARs that were extreme enough to receive +4 ACMG points from computational tools (Table 1). MutPred2 predicted 0 of the possible ECAR substitutions to be extreme enough to receive +4 ACMG points. BayesDel predicted 56 of the 82 possible substitutions to be extreme enough for +4 ACMG points, and these constituted 56 of the 73 such RING and tBRCT +4 results called by BayesDel. AlphaMissense, REVEL, and VEST4 had totals that fell between MutPred2 and BayesDel.

A separate analysis was performed using the BayesDel threshold prescribed in the ENIGMA BRCA1 VCEP (henceforth Bayes-Del VCEP) guidelines (≥ 0.28) as well as for Align-GVGD using C65 as a pathogenic prediction threshold^19^. Under these parameters, which only generate predictions of supporting benign, indeterminate, or supporting pathiogenic, many more of the ECARs identified in the tBRCT domains were predicted to be pathogenic (Table 1). A summary of computational tool performance at ECAR positions and individual scores for each substitution are available in Table S1 and the supplemental spreadsheet.

### Functional Assay Performance

To analyze the performance of functional assays, an OR threshold set to that of the OR of nonsense substitutions observed in our case control dataset for each cancer type was used, which was 16.2 (95% CI: 12.7 – 20.7) for breast cancer and 46.4 (95% CI: 35.7 – 60.7) for ovarian cancer. The combined proportion pathogenic of the breast cancer and ovarian cancer analysis and subsequent ACMG points of functional assay performance using the nonsense substitution OR thresholds are presented in Table 2. ORs and proportions pathogenic by cancer type are presented in the supplemental text. As stated in the Methods section, ACMG points awarded are limited to the maximum number of points outlined in the ACMG guidelines and subsequent transformation to a Bayesian framework.

In the M2H assay, which interrogates variants in the RING domain of BRCA1, variants that were functional and were observed in our case control dataset yielded an adjusted OR close to 1 (1.42, 95% CI: 1.01 – 1.99 for breast cancer and 2.45, 95% CI: 1.41 – 4.24 for ovarian cancer) indicating a group of variants composed almost entirely of benign variants. The combined proportion pathogenic for this group was estimated to be 0.04 which translates to −1 points in the ACMG points system. There were insufficient observations for variants that were categorized as indeterminate to compute an OR or a proportion pathogenic. The group of all variants that were non-functional and observed in our dataset had an adjusted OR of 19.9 (95% CI: 11.3 – 35.0) for breast cancer and 40.0 (95% CI: 21.5 – 73.1) for ovarian cancer which is similar to that of nonsense substitutions for each cancer type in our dataset, indicating a group consisting of mostly pathogenic variants. This analytical subset had a combined proportion pathogenic of 0.99, which would be assigned the full +4 points that are available for non-functional assay results in the ACMG guidelines.

In addition to evaluating the strength of evidence of a single evidence category, our model can also test the strength of combined evidence categories. To this end, the non-functional variants from the M2H assay were stratified into two groups: variants that fell at the ECARs identified from the PMSA (ACMG code: PM1), and all other residues. For variants that were non-functional and fell at an ECAR, the computed OR was 22.0 (95% CI: 12.0 – 40.6) for breast cancer and 40.8 (95% CI: 21.1 – 79.0) for ovarian cancer, slightly higher than the group of all non-functional variants and indicating a subset of variants with similar risk to that of truncating variants. The computed combined proportion pathogenic was 1.00. Combining the +4 points from functional assay and +2 points available from ECARs (PM1), results in +6 points for this group of variants. Thus, using these two evidence criteria alone, these variants reach the Likely Pathogenic threshold.

The computed OR for non-functional variants that fell at other residues was 7.82 (95% CI: 1.68 – 36.3) for breast cancer and 32.9 (95% CI: 6.37 – 170.1) for ovarian cancer, indicating that this group of variants confers less risk than those that are non-functional and fall at an ECAR. The combined proportion pathogenic for the non-functional variants that fell at other residues was 0.70, sufficient for +4 points from functional assay. In the absence of other evidence, this group would remain variants of uncertain significance (VUS).

The SGE assay, which evaluated variants across the RING and tBRCT domains of *BRCA1*, had similar results to the M2H assay. The group of variants that were functional via SGE yielded an OR close to 1 (1.11, 95% CI: 0.91 – 1.36 for breast cancer and 1.34, 95% CI: 0.90 – 2.01 for ovarian cancer) and a combined proportion pathogenic of 0.00, which would qualify for the full −4 points available from a functional assay result. For variants that were non-functional, the SGE assay had an OR of 15.7 (95% CI: 11.1 – 22.0) for breast cancer and 35.1 (95% CI: 24.0 – 51.2) for ovarian cancer and a combined proportion pathogenic of 0.92 which would confer the full +4 points for these variants. Stratification of non-functional variants at ECARs versus non-functional variants at other residues again demonstrates that non-functional variants at ECARs have enough strength of evidence to reach the Likely Pathogenic threshold. The ORs for non-functional variants at ECARs were 23.6 (95% CI: 12.9 – 43.3) for breast cancer and 42.3 (95% CI: 21.9 – 81.7) for ovarian cancer with a combined proportion pathogenic of 1.00, sufficient for +6 ACMG points. The ORs for non-functional variants at non-ECAR positions were 12.2 (95% CI: 8.07 – 18.5) for breast cancer and 31.7 (95% CI: 19.9 – 50.5) for ovarian cancer with a combined proportion pathogenic of 0.76, which qualifies for +4 ACMG points. Moreover, the risk estimates for non-functional variants at ECAR versus non-ECAR positions were distinct from each other, P=0.031 in a two-sided 3×2 FET test.

Whereas the M2H and SGE assays largely confirmed the assumptions of strength of evidence in the ACMG guidelines, the results from the HDR assay in the coiled-coil domain of *BRCA1* produce a different result. The ORs for the functional group of variants were 1.87 (95% CI: 1.11 – 3.15) for breast cancer and 2.04 (95% CI: 0.77 – 5.39) for ovarian cancer with a combined proportion pathogenic of 0.08 translating to 0 ACMG points. The variants that were non-functional via the HDR assay had an OR of 1.58 (95% CI: 0.09 – 26.8) for breast cancer and insufficient observations to calculate an OR for ovarian cancer, with an identical proportion pathogenic of 0.08 and 0 ACMG points. It is noted that there were few observations in our case-control dataset that both fell within this domain and were included in the functional assay. Additionally, many of the variants interrogated by this assay do not appear to be attainable via a single nucleotide substitution to the canonical *BRCA1* transcript, which is the type of substitution that is the primary focus of this analysis.

Due to these limitations, a separate analysis was performed focusing on structurally informed predictions of damage to coiled-coil interactions at the alpha helix spanning amino acids 1,400-1,418, i.e., non-conservative substitutions at the helix *a* and *d* positions plus any substitutions to proline falling in the same interval. With only one observation in a case (p.L1407R) against four control observations (one each for p.M1400K, p.L1404R, p.L1407H, and p.L1414P), the observational evidence weighs against the hypothesis that missense substitutions in this domain are pathogenic (Table 2, final line).

### Combining SGE Functional Results and Computational Predictions

To explore the impact of combining functional assay results and computational tool predictions variants from the SGE assay were stratified by computational prediction (pathogenic, indeterminate, and benign) from seven computational tools (Align-GVGD, AlphaMissense, BayesDel, BayesDel-VCEP, MutPred2, REVEL, and VEST4) using thresholds outlined in Pejaver *et al*.^5, 30^ or those stated in the Methods section. This analysis was performed only on results from the SGE assay which interrogated both the RING and tBRCT domains of *BRCA1* and thus had sufficient observations for stratification. To focus on the impact of combining functional assays and computational tool predictions on variant classification, the combined proportions pathogenic and subsequent ACMG points are presented in Tables 3 and 4. ORs and proportions pathogenic for each stratum and cancer type are presented in the supplemental text (Tables S3 – S6. Multivariate logistic regression was also performed to determine if adding computational tool predictions to functional assay results was statistically significant (Table S7).

### Combining Assay Results of Non-functional with Computational Predictions for Substitutions Falling at Non-ECAR Positions

The proportion pathogenic for the group of variants that were categorized as non-functional via the SGE assay and did not fall at ECARs was 0.76, corresponding to +4 ACMG points, as stated previously. The ORs for this group of variants for breast and ovarian cancer are 12.2 (95% C.I. 8.07 – 18.5) and 31.7 (19.9 – 50.5) respectively. Stratifying these variants by computational prediction allows evaluation of the effect of combining functional assay and computational prediction data toward variant classification (Table 3). It should be noted that if the computational predictions unanimously agreed with the functional assay result, the OR, estimated proportion pathogenic, and ACMG points would remain unchanged after adding in evidence from computational tools. Thus, any utility of computational predictions in this analysis emerges from the ability of the computational tool to accurately disagree with – and thereby stratify – the functional assay result.

For four of the computational tools evaluated, Align-GVGD, AlphaMissense MutPred2, and VEST4, the benign and/or indeterminate prediction yielded a lower proportion pathogenic and lower ACMG point score than the overall pool of SGE non-ECAR, non-functional substitutions. For Align-GVGD and AlphaMissense, this effect of accurate disagreement was strong enough that concordant prediction of pathogenicity from the SGE assay and computational tool resulted in a higher proportion pathogenic and higher ACMG point score (+5). Align-GVGD was the only computational tool that successfully stratified the group of non-functional assay results into three corresponding strata that matched the ACMG expectations. The corresponding ORs for concordant predictions of pathogenicity were 13.9 (95% C.I. 7.39 – 26.1) for breast cancer and 40.2 (95% C.I. 20.1 – 80.3) for ovarian cancer. When Align-GVGD disagreed with the functional assay and predicted variant to be benign the ORs were 6.06 (95% C.I. 1.98 – 18.5) and 22.1 (95% C.I. 6.58 – 74.2) for breast and ovarian cancer respectively.

In contrast, BayesDel and REVEL produced an unexpected reverse stratification, with a higher proportion pathogenic and higher ACMG point score in their indeterminate or benign prediction than in their pathogenic prediction. Correlation between these two - even in failure - is not unexpected as both are meta-callers, incorporating predictions from individual callers into a weighted overall score, and share several of the same individual components ^21, 22^.

### Combining Assay Results of Functional with Computational Predictions for Substitutions Falling at Non-ECAR Positions

For the analysis of variants that were functionally normal via SGE and did not fall at ECARs, an alternative OR ceiling was set at the threshold for moderate-risk variants to evaluate evidence against pathogenicity (OR = 2.0 for breast cancer and 2.9 for ovarian cancer in a general population^8, 9^, which are inflated to 2.31 and 3.83 respectively in our dataset). For the SGE assay, functional variants had an estimated proportion pathogenic of 0.00, sufficient for the −4 ACMG points allowable from functional assays. The OR for breast cancer for this group of variants was 1.11 (95% C.I. 0.91 – 1.36) and 1.34 (95% C.I. 0.90 – 2.01) for ovarian cancer. The results of stratification by computational tool are presented in Table 4; as in Table 3, the utility of computational predictions in this analysis emerges from the ability of a computational tool to accurately disagree with – and thereby stratify – the functional assay result. In this analysis, all of the computational tools were to some extent successful, with their predictions of pathogenicity yielding a higher proportion pathogenic and higher ACMG point score than the overall pool of SGE functional substitutions.

However, BayesDel and REVEL made very few predictions of benignity even though this group of variants were functional in the SGE assay and had ensemble odds ratios and proportions pathogenic consistent with the vast majority actually being benign. On the other hand, for Align-GVGD, AlphaMissense, BayesDel (with and without the VCEP calibration), and VEST4 the effect of discordant prediction of benignity from the SGE assay and pathogenicity by the computational tool was a substantial pool of missense substitutions with an ACMG point score of +1 or higher – though it must be held in mind that these positive scores represent modest evidence in favor of pathogenicity as moderate-risk variants, not as high-risk variants. This is evidenced by the magnitude of the ORs generated for the group of variants predicted to be pathogenic by Align-GVGD: 2.71 (95% C.I. 1.09 – 6.72) for breast cancer and 3.73 (95% C.I. 0.77 – 18.1) for ovarian cancer. Corresponding ORs for the other tools evaluated are presented in the supplemental text (Tables S5 – S6).

Finally, results from multivariate logistic regression substantiate the observed trend in proportions pathogenic and ACMG points provided by computational tool stratification. Predictions from three tools, Align-GVGD, AlphaMissense, and VEST4 were statistically significant (p-values low enough to overcome the intrinsic multiple test) when added to functional assay results. Predictions from BayesDel, BayesDel-VCEP, MutPred2, and REVEL, did not improve the regression model compared to a model that included only the functional assay (Table S7). Note that while adding the results of some computational tools as main effects significantly improved the ability of the model to predict pathogenicity, adding interaction terms between the assay result and the computational tool prediction did not. This indicates that the predictive power of the computational tool is the same regardless of the assay result (wild-type of LOF). Additionally, these logistic regressions did not impose an order on the predictions of the computational tool (pathogenic or benign), however, in general we see that the resulting coefficients reflect our expectations that pathogenic predictions are more indicative of pathogenicity than are benign predictions. Thus, we conclude that Align-GVGD and Alphamissense had the best-balanced performance of variant stratification by providing statistical significance to regression models and meeting the ACMG additivity expectations, as summarized in Table 5.

**Table 5.** Summary of Combining SGE Functional Assay Results with Computational Tool Predictions Compared to ACMG Expectations.

| Computational Tool | Addition of Computational Tool Prediction was Statistically Significant in Logistic Regression Model | Met Expectations When Combined With non-ECAR LOF Results | Met Expectations When Combined With non-ECAR Functionally Normal Results | Met Overall Expectations |
| --- | --- | --- | --- | --- |
| AGVGD | Yes | Yes | Yes | Yes |
| AlphaMissense | Yes | Yes | Yes | Yes |
| BayesDel | No | No | No | No |
| BayesDel – VCEP | Yes | No | Yes | No |
| MutPred2 | No | No | Yes | No |
| REVEL | No | No | No | No |
| VEST4 | Yes | No | Yes | No |

### Example VUS Evaluations

To explore the impact that joint calibration of the SGE functional assay and computational tool prediction might have on variant classification, we work through several examples from ClinVar and/or gnomAD in Table 6. For this analysis, only substitutions with concordant predictions from Align-GVGD and AlphaMissense are considered as these tools performed the best in our analysis thus far. The substitutions selected include one that reaches Likely Pathogenic, three that cover the VUS range from Hot to Cold^38^, and one that reaches Likely Benign. For two of these, p.Asp1840Ala and p.Arg1737Ile, the results from naïve summation of functional and computational points scores would be Likely Benign, whereas the results from joint calibration of functional and computational evidence are VUS.

**Table 6.**
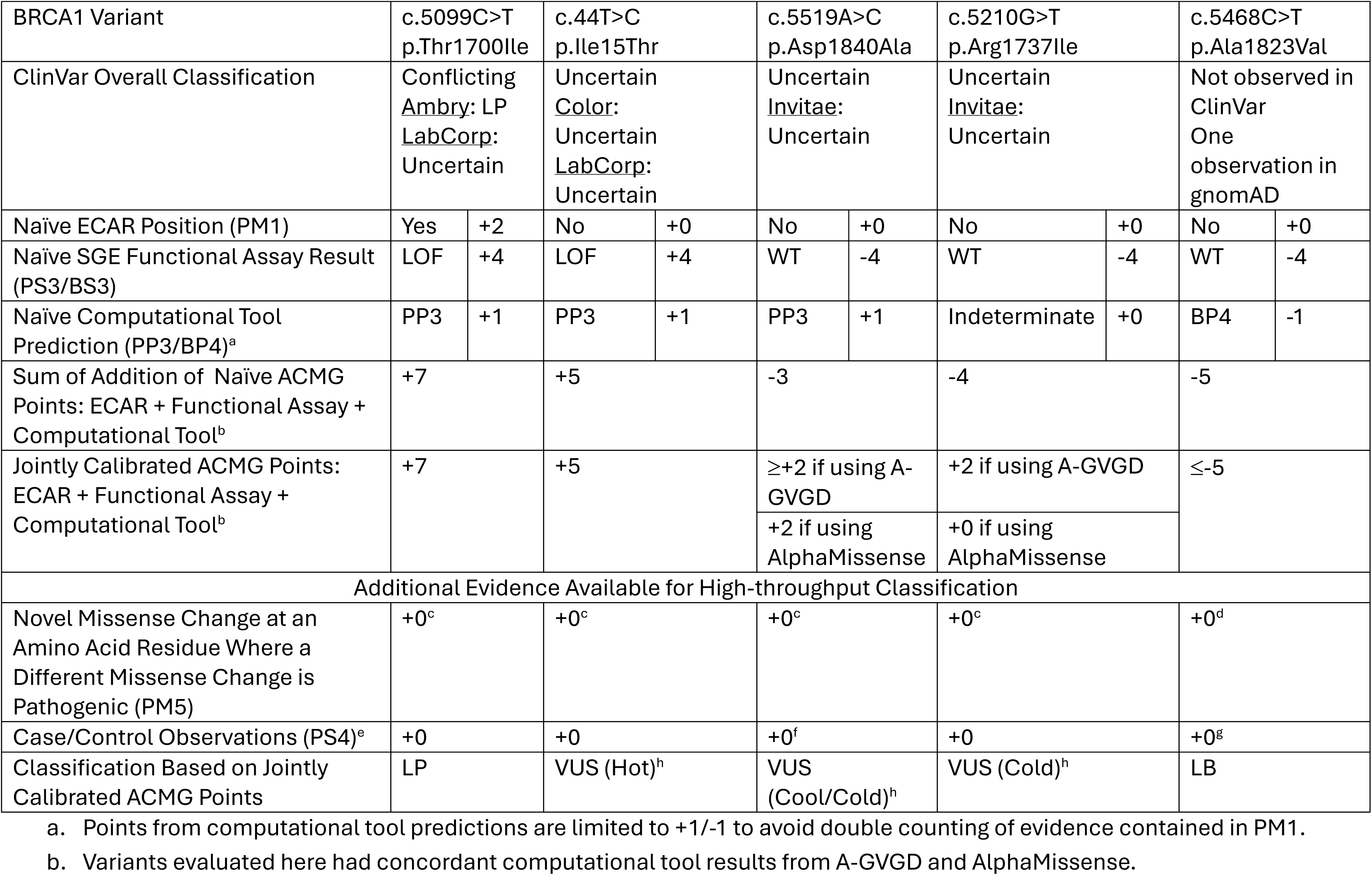
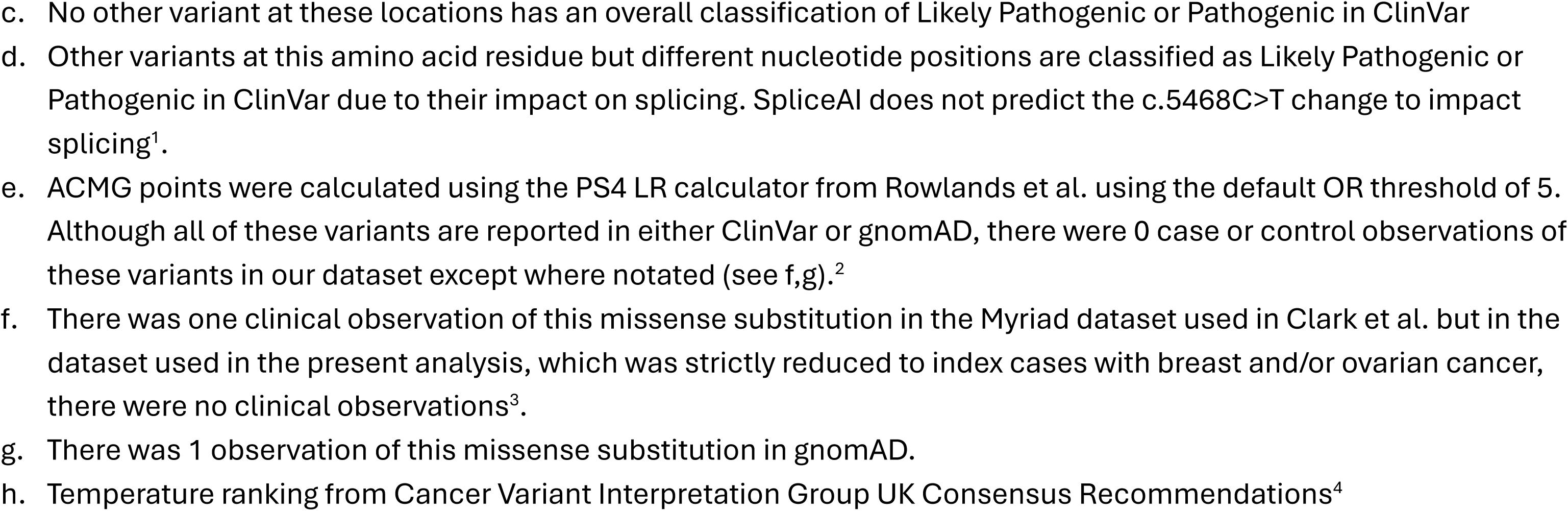
Examples of Di2erences Between Naïve ACMG Addition and Jointly Calibrated Addition of Conserved Domain Data, Functional Assay Results, and Computational Tool Predictions in High-throughput Variant Classification.

|  |  |  |  |  |  |  |  |  |  |  |
| --- | --- | --- | --- | --- | --- | --- | --- | --- | --- | --- |
| BRCA1 Variant | c.5099C>T<br>p.Thr1700Ile |  | c.44T>C<br>p.Ile15Thr |  | c.5519A>C<br>p.Asp1840Ala |  | c.5210G>T<br>p.Arg1737Ile |  | c.5468C>T<br>p.Ala1823Val |  |
| ClinVar Overall Classification | Conflicting<br><u>Ambry</u>: LP<br><u>LabCorp</u>:<br>Uncertain |  | Uncertain<br><u>Color</u>:<br>Uncertain<br><u>LabCorp</u>:<br>Uncertain |  | Uncertain<br><u>Invitae</u>:<br>Uncertain |  | Uncertain<br><u>Invitae</u>:<br>Uncertain |  | Not observed in<br>ClinVar<br>One<br>observation in<br>gnomAD |  |
| Naïve ECAR Position (PM1) | Yes | +2 | No | +0 | No | +0 | No | +0 | No | +0 |
| Naïve SGE Functional Assay Result (PS3/BS3) | LOF | +4 | LOF | +4 | WT | -4 | WT | -4 | WT | -4 |
| Naïve Computational Tool Prediction (PP3/BP4)<sup>a</sup> | PP3 | +1 | PP3 | +1 | PP3 | +1 | Indeterminate | +0 | BP4 | -1 |
| Sum of Addition of Naïve ACMG Points: ECAR + Functional Assay + Computational Tool<sup>b</sup> | +7 |  | +5 |  | -3 |  | -4 |  | -5 |  |
| Jointly Calibrated ACMG Points: ECAR + Functional Assay + Computational Tool<sup>b</sup> | +7 |  | +5 |  | ≥+2 if using A-GVGD |  | +2 if using A-GVGD |  | ≤-5 |  |
|  |  |  |  |  | +2 if using AlphaMissense |  | +0 if using AlphaMissense |  |  |  |
| Additional Evidence Available for High-throughput Classification |  |  |  |  |  |  |  |  |  |  |
| Novel Missense Change at an Amino Acid Residue Where a Different Missense Change is Pathogenic (PM5) | +0<sup>c</sup> |  | +0<sup>c</sup> |  | +0<sup>c</sup> |  | +0<sup>c</sup> |  | +0<sup>d</sup> |  |
| Case/Control Observations (PS4)<sup>e</sup> | +0 |  | +0 |  | +0<sup>f</sup> |  | +0 |  | +0<sup>g</sup> |  |
| Classification Based on Jointly Calibrated ACMG Points | LP |  | VUS (Hot)<sup>h</sup> |  | VUS (Cool/Cold)<sup>h</sup> |  | VUS (Cold)<sup>h</sup> |  | LB |  |

a. Points from computational tool predictions are limited to +1/-1 to avoid double counting of evidence contained in PM1.
b. Variants evaluated here had concordant computational tool results from A-GVGD and AlphaMissense.
- c. No other variant at these locations has an overall classification of Likely Pathogenic or Pathogenic in ClinVar - d. Other variants at this amino acid residue but different nucleotide positions are classified as Likely Pathogenic or Pathogenic in ClinVar due to their impact on splicing. SpliceAI does not predict the c.5468C>T change to impact splicing<sup>1</sup>. - e. ACMG points were calculated using the PS4 LR calculator from Rowlands et al. using the default OR threshold of 5. Although all of these variants are reported in either ClinVar or gnomAD, there were 0 case or control observations of these variants in our dataset except where notated (see f,g).<sup>2</sup> - f. There was one clinical observation of this missense substitution in the Myriad dataset used in Clark et al. but in the dataset used in the present analysis, which was strictly reduced to index cases with breast and/or ovarian cancer, there were no clinical observations<sup>3</sup>. - g. There was 1 observation of this missense substitution in gnomAD. - h. Temperature ranking from Cancer Variant Interpretation Group UK Consensus Recommendations<sup>4</sup>

## DISCUSSION

Here, we derived a maximum likelihood estimate (MLE) model that links the OR for an analytically defined pool of sequence variants within a case-control analysis to an estimate of the proportion of variants within that pool that are pathogenic. Bayes rule is then used to determine the odds of pathogenicity associated with the analysis that defined the pool and thence ACMG points. We also defined Exceptionally Conserved Ancestral Residues (ECARs), a concept that fits within the ACMG PM1 evidence criterion, and then identified ECARs within the RING and tBRCT domains of *BRCA1*. The MLE model was then used to evaluate the strength of evidence for several functional assays alone, in combination with ECAR data, and in combination with computational tool predictions, as summarized in Figure 2. This work takes an important step in the ability to empirically test whether assertions of the strength of evidence for different evidence types, and combinations thereof, proposed in the 2015 ACMG/AMP guidelines are valid. And it does so in a way that takes account of effect size while avoiding circularities inherent in analyses that are based on re-call.

**Figure 2.**
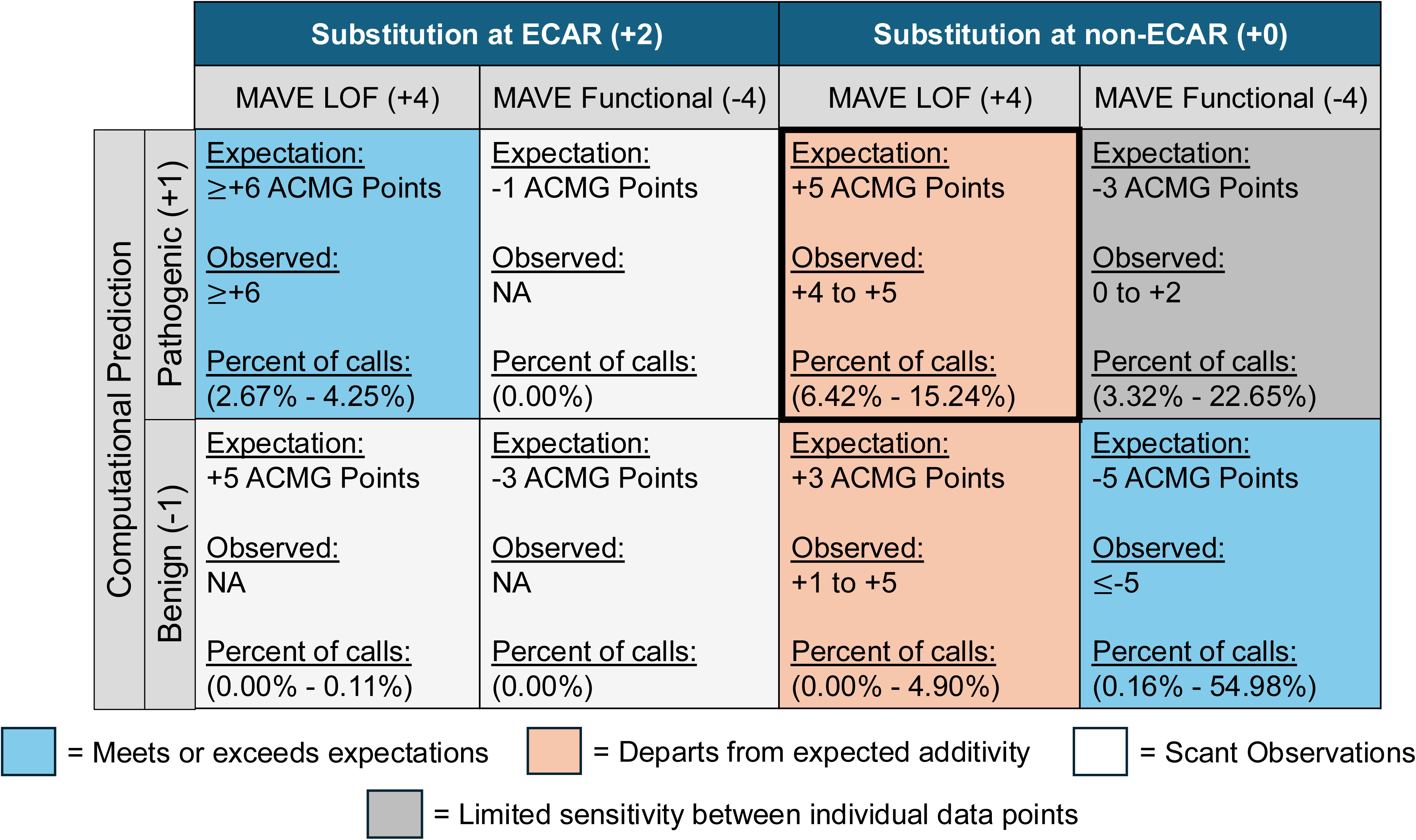
Graphical Summary of Results. The matrix represents the main combinations of evidence evaluated in this study. The number in the row or column headers represent the Bayesian point value assigned to each evidence type in the ACMG guidelines. Each box has three lines: Expectation, Observed, and Percent of calls. “Expectation” is the Bayesian points expected according to the ACMG guidelines. The combination of ECAR, MAVE loss of function, and computational tool pathogenic was capped at ≥+6 points. Otherwise expected points per category are additive as described in the ACMG guidelines. The Observed line represents the Bayesian point value that was observed using the MLE model in this study. The Percent all calls line represents the range, as a percent, of all possible substitutions that were observed for a particular combination of evidence for the computational tools evaluated. Blue filled boxes indicate that the combination of evidence met or exceeded the ACMG guideline expectations in this study. Peach filled boxes indicate that the combination of evidence departed from the expected additivity of the ACMG guideline in this study. The bold black outline highlights that the assumption that MAVE LOF and computational predictions of pathogenicity are additive is not met in this study; moreover, this combination had a substantial number of observations. White filled boxes represent combinations of evidence with little to no data. Dark grey filled boxes represent combinations of evidence with results that may be beyond the dynamic range of the MLE model.

Results from three *BRCA1* functional assays were examined: a saturation genome editing (SGE) assay that examined both the RING and tBRCT domain, a mammalian 2-hybrid (M2H) assay that examined the RING domain, and a homology directed repair (HDR) assay that examined the coiled-coil domain through which BRCA1 interacts with PALB2. With enormous differences in the case-control ORs calculated for non-functional versus functional missense substitutions, the SGE and M2H assays both provide significant evidence towards sequence variant classification, which will be discussed in greater detail below. In contrast, there was no OR-based evidence that loss of function in the coiled-coil HDR assay predicted increase risk of breast or ovarian cancer; moreover, null results from a structurally informed analysis of predicted loss of function substitutions in the core alpha helix of the coiled-coil domain (amino acids 1,400 – 1,418) weighs against the hypothesis that missense substitutions in this domain can be pathogenic. Therefore, careful evaluation of functional assays is required prior to their use in clinical settings and +/− 4 ACMG points should not be assigned carte blanche^6^.

Using an evolutionarily deep multi-supergroup protein multiple sequence alignment of BRCA1, BARD1, and related RING-tBRCT proteins, we identified 14 ECARs in *BRCA1* (and *BARD1*). Stratification of ECAR substitutions by *BRCA1* MAVE functional assay results, followed by analysis through the MLE model, led to three interesting observations. (1) Of 82 possible missense substitutions at an ECAR, 78 had loss of function in the SGE; the remaining four were either indeterminate or not assessed by the SGE assay, and 2 of these had loss of function in the M2H assay. (2) Loss of function ECAR substitutions had an OR very similar to that of *BRCA1* nonsense substitutions; this is an example of the expected additivity between evidence criteria, consistent with +6 ACMG points for the combination. (3) The 326 non-functional substitutions at non-ECAR positions also have an elevated OR, but significantly lower than that observed for substitutions at ECAR positions, corresponding to +4 points.

With the recent publication of ACMG points-based calibration of several computational tools, including AlphaMissense, it is of interest to examine results from combining computational tool evidence with functional assay evidence. Since ECARs fit within the PM1 evidence criterion, yet should ideally be captured by computational tools, we avoided double counting ECAR evidence by applying the computational tools to functional assay results at the non-ECAR RING and tBRCT positions.

For the set of 326 non-ECAR RING and tBRCT substitutions with SGE assay loss-of-function, the expected result is that computational tool evidence in favor of pathogenicity will result in an analytic subset with a higher OR, adding an ACMG point(s) to the score of the input data set. Computational tool evidence in favor of benignity should then result in an analytic subset with a lower OR, subtracting an ACMG point(s) from the score of the input data set. Of the six computational tools tested, only two – Align-GVGD and AlphaMissense – generated this bidirectional pattern. Two other tools – MutPred2 and VEST4 – generated negative points with their indeterminate and/or evidence of benignity outputs, but did not generate positive points with their evidence of pathogenicity. It should be noted that for these non-ECARs substitutions, concordant evidence of pathogenicity from the functional assay and either Align-GVGD or AlphaMissense resulted in a score of +5 points, meaning that additional evidence in favor of pathogenicity – perhaps from co-segregation or tumor features as was the case with *BRCA1* p.Pro34Ser^7^ – would be required for the variants to reach ≥ +6 points and thus Likely Pathogenic. Under the computational tool score thresholds established in Pejvaer et al.^5, 30^ for Pathogenic Supporting (+1 ACMG point), most tools added 0 ACMG points when combined with loss of function results (except for AlphaMissense). When these thresholds were changed to that of Pathogenic Moderate (+2 ACMG points), no tool added +2 ACMG points on top of the functional assay result (Table S8), which is another example of departure from simple additivity assumed in the ACMG guidelines.

Under the logic of both the IARC/ENIGMA/InSiGHT and the Bayesian interpretation of the ACMG/AMP sequence variant classification systems, evidence against pathogenicity has been interpreted as evidence in favor of benignity. At the same time, both ENIGMA (Evidence-based Network for the Interpretation of Germline Mutant Alleles) and the BRCA1/2 VCEP (Variant Classification Expert Panel) recognize the existence of moderate-risk susceptibility genes and moderate-risk variants of high-risk susceptibility genes (also called Reduced Penetrance Pathogenic Variants, RPPV)^8, 19, 39^. To reduce the likelihood of generating evidence in favor of benignity for missense substitutions that might eventually be classified as RPPVs, we made the potentially controversial choice to estimate proportions pathogenic and ACMG points for variants with normal function in the functional assays using moderate-risk breast and ovarian cancer thresholds.

For the set of 1,284 RING and tBRCT missense substitutions that were functional in the SGE assay, the ORs for breast and ovarian cancer were near 1.0, consistent with −4 ACMG points, even if calculated with the moderate-risk threshold. Stratification of these with the set of six computational tools revealed two trends plus an analytic difficulty. First, two of the computational tools – BayesDel and REVEL – returned scores in favor of benignity for only a very small fraction of these missense substitutions: 3% by BayesDel and <1% by REVEL. We interpret this as a failure within these two tools’ algorithms. Second, three of the tools – Align-GVGD, AlphaMissense, and VEST4 – generated a bidirectional stratification, with higher ORs and ACMG point scores associated with computational evidence of pathogenicity and lower ORs and point scores associated with evidence of benignity. We interpret this as a subset of the computational tools adding useful evidence towards variant classification on top of the functional assay result. The analytic difficulty is that modest changes in the OR produced large changes in the ACMG point scores; when the SGE assay and computational tool were discordant, combining the two produced point scores that were beyond the range expected from simple additivity.

The individual sequence variants evaluated in Table 6 provide examples for many of the trends described above. For example, functional assay loss of function at an ECAR, with concordant computational evidence, (p.Thr1700Ile) results in Likely Pathogenic. On the other hand, loss of function at a non-ECAR position with concordant computational evidence in favor of pathogenicity (p.Ile15Thr) results in a hot VUS, and further evidence of pathogenicity would be required to reach Likely Pathogenic. In either scenario, evidence in favor of benignity that might arise from human subjects data or other evidence could move variants with these evidence combinations from Likely Pathogenic and hot VUS to hot or cool VUS respectively. At the other end of the spectrum, and using the moderate-risk OR threshold for calibration to reduce the likelihood of classifying RPPVs as likely benign, an assay result of functional with concordant computational evidence of benignity (p.Ala1823Val) results in Likely Benign. But discordant results between a calibrated functional assay and calibrated computational tool (p.Asp1840Ala, p.Arg1737Ile) result in cool to cold VUS – and a concern that the computational tools need to be accurately calibrated in order to avoid this classification result for too many missense substitutions.

Like most studies, this one had several limitations. A challenge to the ECARs component of the approach pursued here is variable quality of assembly and subsequent annotation of reference genomes. Thus, searches against available databases may give a misleading view of which organisms outside of Holozoa encode genes with both RING and tBRCT domains. Furthermore, this study was conducted on *BRCA1*, a gene that probably shares ancestry with *BARD1*, these two representing Holozoan examples of a gene family that has undergone a modest level of evolutionary diversification. It is not known if this method will be applicable to genes which are members of gene families that have undergone wide diversification or that are unique to Holozoa. Much of the subsequent results depend on case-control estimates of OR and downstream calculations. However, carrier data were too sparse, and the racial/ethnic categorizations between our source data too different for us to make detailed adjustment for race/ethnicity, let alone other potentially relevant variables. These concerns may impact the precision of our results, especially at effect sizes that are relevant to distinctions between benign and moderate-risk missense substitutions. Finally, repeated use of the MLE model, especially applying it at two distinctly different OR thresholds, may have overstressed its assumption of a 2-component mixture. This concern may also impact the precision of our results.

The MLE model coupled with a preliminary definition of ECARs allowed us to tease out the observation that substitutions falling at the 14 *BRCA1*/*BARD1* ECAR residues contribute disproportionate evidence of pathogenicity for missense substitutions in *BRCA1*, leading to a hypothesis that will need to be tested with evidence from other genes. With substitutions falling at ECAR positions set aside, Align-GVGD and AlphaMissense emerged as the two strongest computational tools for prediction of *BRCA1* missense substitution pathogenicity. These two tools also performed well for prediction of *BRCA1* missense substitution benignity. Since AlphaMissense is a proteome-wide tool whereas Align-GVGD is not, this observation leads to a recommendation in favor of the use of AlphaMissense – but also a recommendation that one of the Meta-predictor computational tools be upgraded to include explicit recognition of ECARs, an Align-GVGD like scoring component, and AlphaMissense itself.

The MLE model also enabled joint evaluation of functional assay and computational tool outputs with separate OR thresholds for evidence in favor of pathogenicity and evidence in favor of benignity. In both instances, discordant results from functional assay and computational tool created analytical subsets of variants that may harbor variants that are actually RPPV, which otherwise may have been incorrectly classified as high-risk Likely Pathogenic or Likely Benign. On the benign end of the spectrum, this analysis was only possible because the BRCA1/2 VCEP has a well-defined moderate-risk threshold. As the findings from this study are applied to other health outcomes, a critical effort will be defining similar moderate-risk thresholds for the relevant set of genes and diseases. Doing so will enable the analytical detection of the biological reality that disease risk is not an all or nothing outcome but rather falls on a continuum from benign to pathogenic.

## Data Availability

Simple observational data tables are available from S.V.T. R code for the main analysis is available from A.T.

## ACKNOWLEDGEMENTS

We would like to thank Marcy Richardson, Tina Pesaran, Colin Young, and Steven Harrison from Ambry Genetics for thoughtful discussion and database support. We would also like to thank AC Tan, Jay Gertz, and Pinar Bayrak-Toydemir for thoughtful reading and comments on the manuscript. This work was funded in part by NIH NCI R01CA264971, to SVT. STP is a recipient of the NIH NCI T32CA265782.

## DECLARATION OF INTERESTS

The authors declare no competing interests.

## Notes

### Competing Interest Statement

The authors have declared no competing interest.

### Funding Statement

This work was funded in part by NIH NCI R01CA264971, to SVT. STP is a recipient of NIH NCI T32CA265782.

### Author Declarations

The ethics committee IRB of the University of Utah has decided that the research described in this study is not human subjects research and thus did not require any further IRB approval.

### Summary of Updates

Analysis was conducted separately for breast and ovarian cancer and the results were then combined to evaluate joint functional assay and computational tool performance.

