## Supplemental Figures and Tables for "When Two Plus Four Does Not Equal Six: Combining Computational and Functional Evidence to Classify BRCA1 Key Domain Missense Substitutions"

Supplemental Figure 1

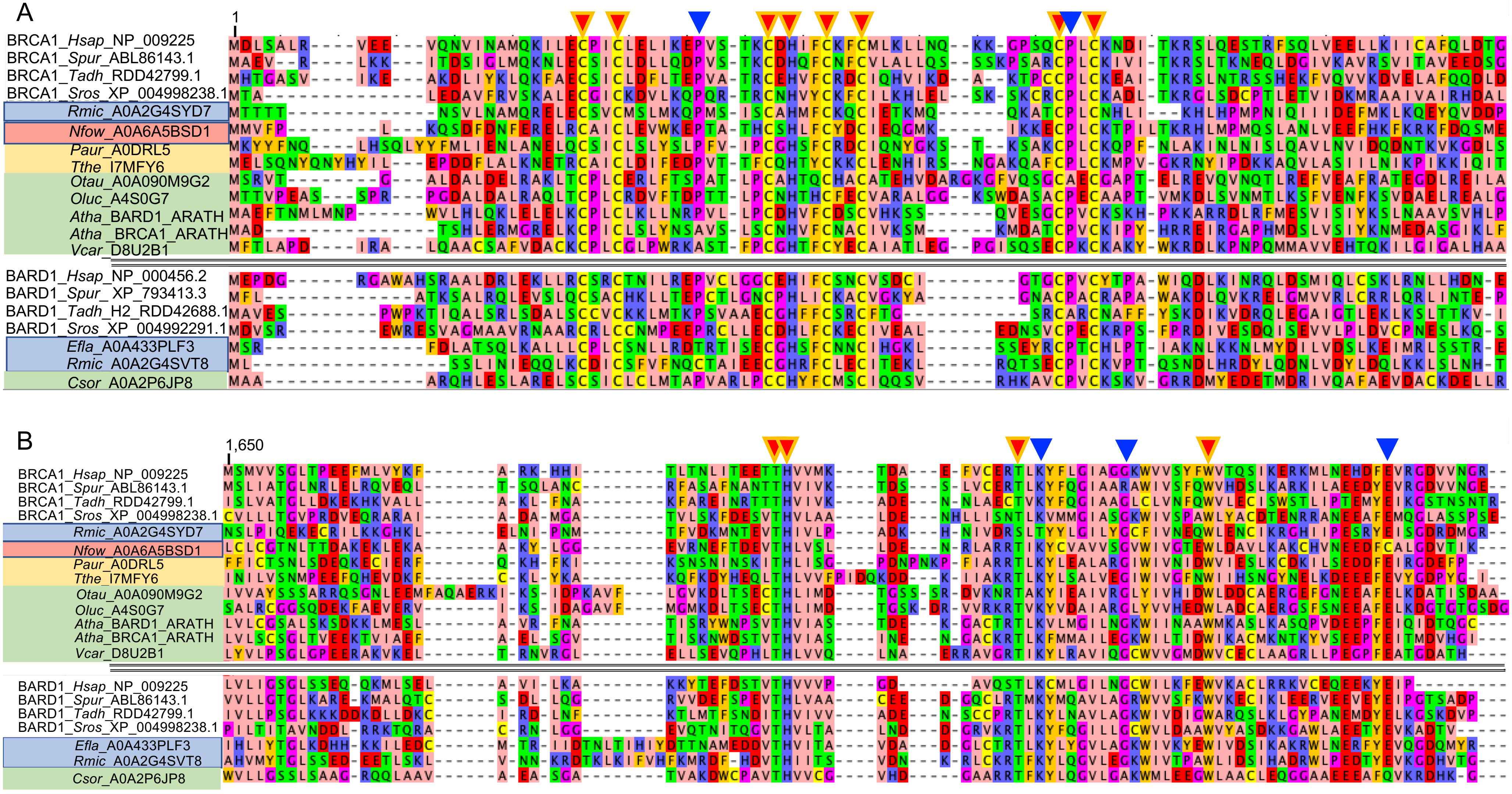

Supplemental Figure 1

C

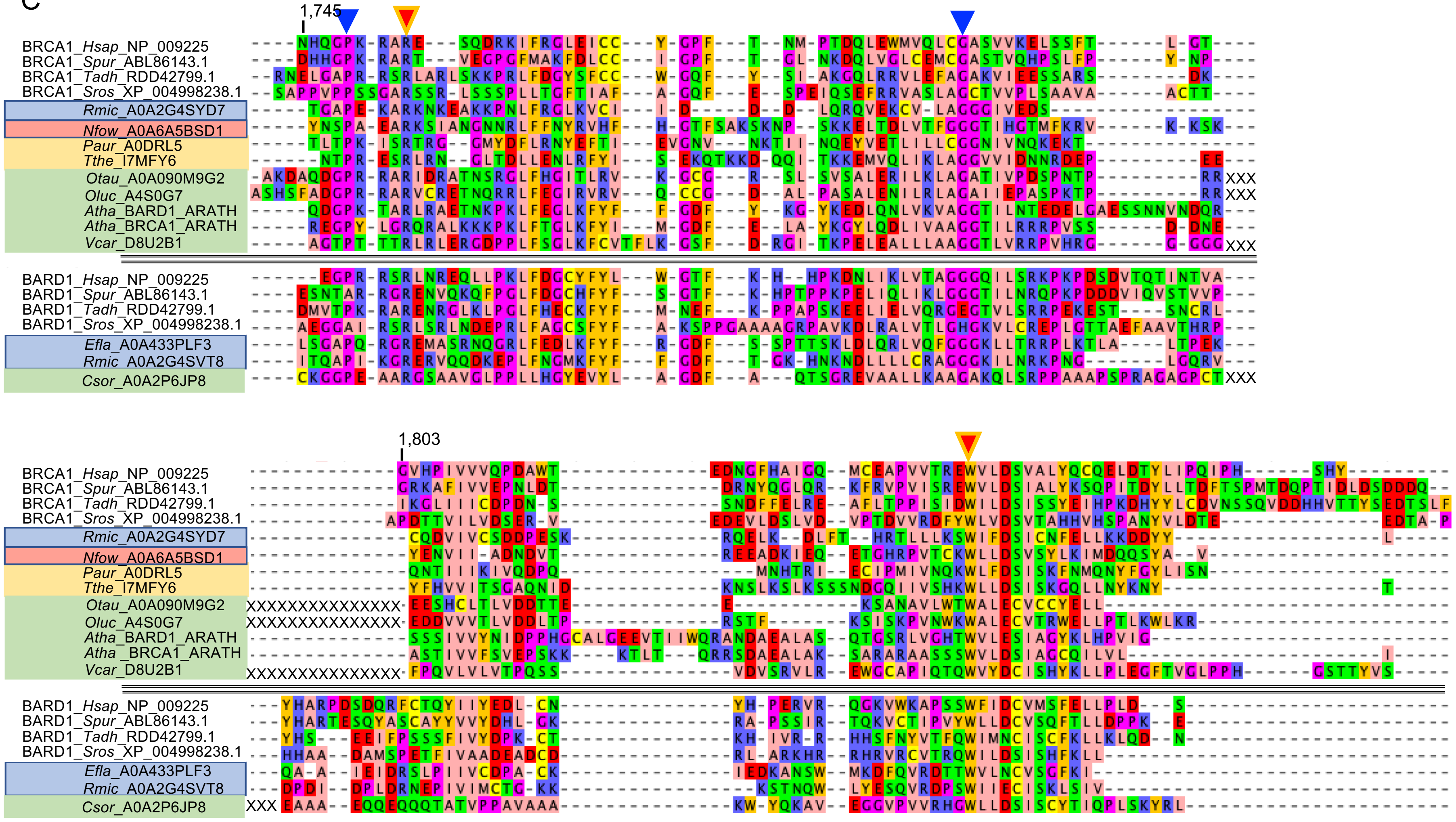

**Supplemental Figure 1**. Multi-Supergroup RING-tBRCT Protein Multiple Sequence Alignment and Selection of Exceptionally Conserved Ancestral Residues (ECARs).

Figure annotations. Plain background: Holozoa (Animals). *Hsap*, *Homo sapiens*; *Spur*, *Strongylocentrotus purpuratus*; *Tadh*, *Trichoplax adhearans*; *Sros*, *Salpingoeca rosetta*. Blue background: Holomycota (Fungi). *Efla*, Endogone FLAS-F59071; *Rmic*, *Rhizopus microsporus*. Red background: Discoba. *Nfow*, *Naeglaeria fowleri*. Beige background: SAR (Stramenopila / Alveolata / Rhizaria). *Paur*, *Paramecium aurelia*; *Tthe*, *Tetrahymena thermophila*. Green background: Archaeplastida (Plants). *Otau*, *Ostreococcus tauri*; *Oluc*, *Ostreococcus lucimarus*; *Atha*, *Arabidopsis thaliana*; *Vcar*, *Volvox carteri*; *Csor*, *Chlorella sorokiniana*. Above the double divider line: *BRCA1*, *BRCA1*-like, and ambiguously related RING-tBRCT gene sequences. Below the double divider line: *BARD1* and *BARD1*-like RING-tBRCT gene sequences. Orange-red arrows: ECAR residues. Blue arrows: well conserved positions that are nonetheless excluded from the ECAR set. “XXXXX”: Several of the Archaeplastida RING-tBRCT sequences have low sequence complexity insertions at a position corresponding to the end of exon 22 of human *BRCA1*. These have been removed and replaced with “XXX” for clarity; indeed, it was difficult to prepare clean alignments of the second half of BRCT2 before these insertions were recognized.

**A**. RING domain alignment. Using human *BRCA1* numbering, the ECAR residues in this domain are: Cys24, Cys27, Cys39, His41, Cys44, Cys47, Cys61, and Cys64. Met1 is not included in the list because the mechanism of loss-of-function for translation initiator substitutions is different than for open reading frame missense substitutions. The next two best conserved positions are Pro34 and Pro62; however, they are excluded from the ECAR list because the positions do not reach the level of conservation that we required (no alternate amino acids among metazoan sequences included in the alignment, <10% alternate amino acids, all conservative with respect to the ancestral residue, in the included sequences from other supergoups). Note that the first amino acid included in the human *BARD1* sequence is actually Met26.

**B**. Alignment of BRCT1. Using human *BRCA1* numbering, the ECAR residues in this domain are : Thr1685, His1686, Thr1700, and Trp1718. The next three best conserved positions are Lys1702, Gly1710 and Glu1735; however, they are excluded from the ECAR list because the positions do not reach the level of conservation that we required (No alternate amino acids among metazoan sequences included in the alignment. <10% alternate amino acids, all conservative with respect to the ancestral residue, in the included sequences from other supergoups).

**C**. Alignment of BRCT2. Using human *BRCA1* numbering, the ECAR residues in this domain are: Arg1753 and Trp1837. The next two best conserved positions are Pro1749 and Gly1788; however, they are excluded from the ECAR list because the positions do not reach the level of conservation that we required (No alternate amino acids among metazoan sequences included in the alignment. <10% alternate amino acids, all conservative with respect to the ancestral residue, in the included sequences from other supergoups).

**Gene family notes**. (**1**) An Important point is that the Holomycota / *Rhizopus microsporus* sequence A0A2G4SYD7, the Discoba / *Naeglaeria fowleri* sequence A0A6A5BSD1, and the Alveolata / *Paramecium aur*elia & *Tetrahymena thermophila* sequences A0DRL5 & I7MFY6 are all small proteins with only a few amino acids between their RING and tandem BRCT domains. A reasonable hypothesis would be that these resemble the ancestral state of the RING-tBRCT gene family. (**2**) In the Holomycota / Mucoromycota, we unexpectedly find genomes that encode two distinct RING-tBRCT gene from structurally distinct subfamilies, e.g., *Rhizopus microsporus*. One (A0A2G4SYD7) encodes a small protein with RING and tBRCT sequences that resemble those of Holozoan *BRCA1*. The other (A0A2G4SVT) has the domain structure RING-RanBP Zn finger-tBRCT. (**3**) In the Archaeplastida, we unexpectedly even greater RING-tBRCT subfamily complexity, e.g., RING-tBRCT (*Volvox carteri* D8U2B1_VOLCA)†, RING-Extended PHD-tBRCT (*Arabidopsis thaliana* BRCA1_ARATH and BARD1_ARATH)†, RING-RanBP Zn finger-tBRCT (*Chlorella sorokiniana* A0A2P6TJP8_CHLSO), and RING-coil-tBRCT (*Ostreococcus tauri* A0A090M9G2_OSTTA). †Note that these two combinations were described by Trapp et al, 2011, PMID: 22629260, main text citation #30. (**4**) Overall, the question of when *BRCA1* and *BARD1* diverged from an ancestral gene remains, to our knowledge, unanswered.

**Table S1. Breakdown of Computational Predictions at Exceptionally Conserved Ancestral Residues (ECAR)**

| Computational Tool | BP4[a] | Indeterminate[b] | PP3(+1,+2)[c] | PP3(+4)[d] | NA[e] | Total |
|---|---|---|---|---|---|---|
| A-GVGD | 0 | 14 | 68 | 0[f] | 0 | 82 |
| AlphaMissense | 1 | 4 | 36 | 41 | 0 | 82 |
| BayesDel | 0 | 0 | 26 | 56 | 0 | 82 |
| BayesDel-VCEP | 2 | 1 | 79 | 0 | 0 | 82 |
| MutPred2 | 1 | 28 | 52 | 0[f] | 1 | 82 |
| REVEL | 0 | 0 | 48 | 34 | 0 | 82 |
| VEST4 | 0 | 2 | 68 | 12 | 0 | 82 |

a. Predicted benign according to thresholds established in the Methods section.
b. These variants have scores that do not fall in the predicted benign or predicted pathogenic ranges.
c. Predicted pathogenic with a score sufficient for either +1 or +2 ACMG point(s) according to the calibration performed by Pejaver et al.
d. Predicted pathogenic with a score sufficient for +4 ACMG points according to the calibration performed by Pejaver et al.
e. A computational score was not available for these variants.
f. A-GVGD and BayesDel-VCEP were not calibrated for +4 ACMG points and thus all pathogenic predictions from these tools are in the column labeled PP3(+1,+2).

**Table S2. Odds Ratios and Proportions Pathogenic by Cancer for each Functional Assay Result.**

| Assay Result | Control Obs. | Case Obs. | Odds Ratio[a] | 95% C.I | Proportion Pathogenic |
|---|---|---|---|---|---|
| M2H Assay – Breast Cancer | | | | | |
| Functional | 75 | 65 | 1.42[b] | 1.01-1.99 | 0.05 |
| Indeterminate | 0 | 1 | NA | NA | NA |
| Non-functional All | 13 | 187 | 19.9[b] | 11.3-35.0 | 1 |
| Non-functional ECAR | 11 | 177 | 22.0 | 12.0-40.6 | 1 |
| Non-functional Other | 2 | 10 | 7.82 | 1.68-36.3 | 0.67 |
| Nonsense | 70 | 765 | 16.2 | 12.7-20.8 | Reference (1) |
| M2H Assay – Ovarian Cancer | | | | | |
| Functional | 75 | 16 | 2.45[b] | 1.41-4.24 | 0.04 |
| Indeterminate | 0 | 0 | NA | NA | NA |
| Non-functional All | 13 | 58 | 40.0[b] | 21.5-73.1 | 0.98 |
| Non-functional ECAR | 11 | 52 | 40.8 | 21.1-79.0 | 1.00 |
| Non-functional Other | 2 | 6 | 32.9 | 6.37-170.1 | 0.73 |
| Nonsense | 70 | 314 | 46.4 | 35.7-60.7 | Reference (1) |
| SGE Assay – Breast Cancer | | | | | |
| Functional | 239 | 162 | 1.11[b] | 0.91-1.36 | 0.00 |
| Indeterminate | 24 | 29 | 2.17 | 1.25-3.77 | 0.13 |
| Non-functional All | 37 | 371 | 15.7[b] | 11.1-22.0 | 0.96 |
| Non-functional ECAR | 11 | 189 | 23.6 | 12.9-43.3 | 1.00 |
| Non-functional Other | 26 | 182 | 12.2 | 8.07-18.5 | 0.82 |
| Nonsense | 70 | 765 | 16.2 | 12.7-20.8 | Reference (1) |
| SGE Assay – Ovarian Cancer | | | | | |
| Functional | 239 | 27 | 1.34[b] | 0.90-2.01 | 0.00 |

| | | | | | |
|---|---|---|---|---|---|
| Indeterminate | 24 | 10 | 5.29 | 2.47-11.3 | 0.10 |
| Non-functional All | 37 | 122 | 35.1[b] | 24.0-51.2 | 0.79 |
| Non-functional ECAR | 11 | 54 | 42.3 | 21.9-81.7 | 1.00 |
| Non-functional Other | 26 | 68 | 31.7 | 19.9-50.5 | 0.66 |
| Nonsense | 70 | 314 | 46.4 | 35.7-60.7 | Reference (1) |
| HDR Assay – Breast Cancer | | | | | |
| Functional | 27 | 33 | 1.87 | 1.11-3.15 | 0.14 |
| Indeterminate | 15 | 0 | NA | NA | NA |
| Non-functional All | 1 | 1 | 1.58 | 0.09-26.8 | 0.08 |
| A priori damaging[c] | 4 | 1 | 0.46 | 0.50-4.24 | 0.00 |
| Nonsense | 70 | 765 | 16.2 | 12.7-20.8 | Reference (1) |
| HDR Assay – Ovarian Cancer | | | | | |
| Functional | 27 | 5 | 2.04 | 0.77-5.39 | 0.03 |
| Indeterminate | 15 | 0 | NA | NA | NA |
| Non-functional All | 1 | 0 | NA | NA | NA |
| A priori damanging[c] | 4 | 0 | NA | NA | NA |
| Nonsense | 70 | 314 | 46.4 | 35.7-60.7 | Reference (1) |

a. Adjusted for race/ethnicity

b. These ORs are statistically different from one another with a p-value less than 0.0001.

c. This subset interrogates the substitutions to Proline and other non-conservative amino acid substitutions that fell at the a,d positions of the alpha helix in the coiled-coil domain (M1400, Q1401, H1402, L1404, L1407, Q1408, Q1409, M1411, A1412, L1414, A1416, L1418), regardless of whether they were interrogated in the assay. Only Q1401P was included in the assay and had no clinical or gnomAD observations in our dataset.

**Table S3. Odds Ratios and Proportions Pathogenic for Breast Cancer After Computational Tool Stratification of Loss of Function Results from SGE Assay**

| Computational Tool Prediction | Control Obs. | Case Obs | Odds Ratio[a] | 95% C.I. | Proportion Pathogenic |
|---|---|---|---|---|---|
| AGVGD | | | | | |
| Pathogenic | 11 | 87 | 13.9 | 7.39-26.1 | 0.87 |
| Indeterminate | 11 | 77 | 12.5 | 6.65-23.7 | 0.82 |
| Benign | 4 | 15 | 6.06 | 1.98-18.5 | 0.55 |
| AlphaMissense | | | | | |
| Pathogenic | 19 | 167 | 15.4 | 9.54-24.8 | 0.91 |
| Indeterminate | 7 | 8 | 1.86 | 0.66-5.24 | 0.12 |
| Benign | 0 | 1 | NA | NA | NA |
| BayesDel | | | | | |
| Pathogenic | 25 | 173 | 12.0 | 7.86-18.3 | 0.81 |
| Indeterminate | 1 | 9 | 15.1 | 1.89-120.9 | 0.92 |
| Benign | 0 | 0 | NA | NA | NA |
| BayesDel-VCEP | | | | | |
| Pathogenic | 23 | 165 | 12.5 | 8.06-19.4 | 0.83 |
| Indeterminate | 2 | 7 | 5.51 | 1.12-27.1 | 0.52 |
| Benign | 1 | 10 | 16.2 | 2.05-128.3 | 0.96 |
| MutPred2 | | | | | |
| Pathogenic | 15 | 110 | 13.3 | 7.70-22.8 | 0.84 |
| Indeterminate | 10 | 70 | 11.3 | 5.77-22.0 | 0.82 |
| Benign | 1 | 2 | 3.70 | 0.32-42.2 | 0.30 |
| REVEL | | | | | |
| Pathogenic | 25 | 170 | 11.8 | 7.73-18.0 | 0.81 |
| Indeterminate | 1 | 12 | 19.6 | 2.53-152.2 | 1.00 |
| Benign | 0 | 0 | NA | NA | NA |
| VEST4 | | | | | |
| Pathogenic | 21 | 164 | 13.5 | 8.51-21.3 | 0.87 |
| Indeterminate | 5 | 18 | 6.46 | 2.37-17.6 | 0.53 |
| Benign | 0 | 0 | NA | NA | NA |

a. Adjusted for race/ethnicity.

**Table S4. Odds Ratios and Proportions Pathogenic for Ovarian Cancer After Computational Tool Stratification of Loss of Function Results from SGE Assay**

| Computational Tool Prediction | Control Obs. | Case Obs | Odds Ratio[a] | 95% C.I. | Proportion Pathogenic |
|---|---|---|---|---|---|
| A-GVGD | | | | | |
| Pathogenic | 11 | 36 | 40.2 | 20.1-80.3 | 0.78 |
| Indeterminate | 11 | 23 | 25.4 | 12.1-53.1 | 0.54 |
| Benign | 4 | 9 | 22.1 | 6.58-74.2 | 0.58 |
| AlphaMissense | | | | | |
| Pathogenic | 19 | 53 | 34.4 | 20.1-58.8 | 0.69 |
| Indeterminate | 7 | 7 | 9.87 | 3.35-29.1 | 0.27 |
| Benign | 0 | 2 | NA | NA | NA |
| BayesDel | | | | | |
| Pathogenic | 25 | 63 | 30.1 | 18.7-48.5 | 0.64 |
| Indeterminate | 1 | 5 | 52.7 | 5.89-470.4 | 1.00 |
| Benign | 0 | 0 | NA | NA | NA |
| BayesDel-VCEP | | | | | |
| Pathogenic | 23 | 61 | 32.6 | 19.9-53.4 | 0.67 |
| Indeterminate | 2 | 2 | 6.83 | 0.96-48.5 | 0.27 |
| Benign | 1 | 5 | 52.7 | 5.89-470.4 | 1.00 |
| MutPred2 | | | | | |
| Pathogenic | 15 | 42 | 31.7 | 17.3-58.1 | 0.70 |
| Indeterminate | 10 | 25 | 32.0 | 15.1-68.0 | 0.63 |
| Benign | 1 | 1 | 11.5 | 0.66-203.1 | 0.27 |
| REVEL | | | | | |
| Pathogenic | 25 | 62 | 29.8 | 18.5-48.1 | 0.63 |
| Indeterminate | 1 | 6 | 60.1 | 6.98-518.2 | 1.00 |
| Benign | 0 | 0 | NA | NA | NA |
| VEST4 | | | | | |
| Pathogenic | 21 | 58 | 33.8 | 20.2-56.4 | 0.69 |
| Indeterminate | 5 | 10 | 20.1 | 6.65-60.7 | 0.52 |
| Benign | 0 | 0 | NA | NA | NA |

a. Adjusted for race/ethnicity.

**Table S5. Odds Ratios and Proportions Pathogenic for Breast Cancer After Computational Tool Stratification of Wild-type Results from SGE Assay**

| Computational Tool Prediction | Control Obs. | Cases Obs. | Odds Ratio[a] | 95% C.I. | Proportion Pathogenic[b] |
|---|---|---|---|---|---|
| AGVGD | | | | | |
| Pathogenic | 8 | 12 | 2.71[c] | 1.09-6.72 | 0.92 |
| Indeterminate | 43 | 47 | 1.76 | 1.15-2.68 | 0.49 |
| Benign | 188 | 103 | 0.88[c] | 0.69-1.13 | 0.00 |
| AlphaMissense | | | | | |
| Pathogenic | 17 | 22 | 2.12[c] | 1.11-4.05 | 0.71 |
| Indeterminate | 115 | 93 | 1.31 | 0.99-1.74 | 0.16 |
| Benign | 107 | 47 | 0.71[c] | 0.50-1.01 | 0.00 |
| BayesDel | | | | | |
| Pathogenic | 77 | 79 | 1.69 | 1.22-2.32 | 0.42 |
| Indeterminate | 155 | 76 | 0.79 | 0.60-1.04 | 0.00 |
| Benign | 7 | 7 | 1.67 | 0.57-4.86 | 0.39 |
| BayesDel-VCEP | | | | | |
| Pathogenic | 28 | 27 | 1.53 | 0.89-2.62 | 0.34 |
| Indeterminate | 45 | 44 | 1.59 | 1.04-2.43 | 0.36 |
| Benign | 166 | 91 | 0.89 | 0.69-1.16 | 0.00 |
| MutPred2 | | | | | |
| Pathogenic | 7 | 5 | 1.46 | 0.46-4.67 | 0.04 |
| Indeterminate | 79 | 70 | 1.35 | 0.97-1.87 | 0.25 |
| Benign | 153 | 87 | 0.95 | 0.73-1.24 | 0.00 |
| REVEL | | | | | |
| Pathogenic | 85 | 71 | 1.38 | 1.00-1.91 | 0.19 |
| Indeterminate | 154 | 91 | 0.95 | 0.73-1.23 | 0.00 |
| Benign | 0 | 0 | NA | NA | NA |
| VEST4 | | | | | |
| Pathogenic | 15 | 27 | 3.43[c] | 1.81-6.51 | 1.00 |
| Indeterminate | 139 | 108 | 1.18 | 0.91-1.52 | 0.12 |
| Benign | 85 | 27 | 0.57[c] | 0.36-0.88 | 0.00 |

a. Adjusted for race/ethnicity.
b. The OR threshold used for this calculation is that of moderate risk variants (OR = 2 for breast cancer and OR = 2.9 for ovarian cancer) adjusted for observational inflation in our data to OR = 2.31 and OR = 3.83 respectively.
c. Statistically significant with a p-value less than 0.05.

**Table S6. Odds Ratios and Proportions Pathogenic for Ovarian Cancer After Computational Tool Stratification of Wild-Type Results from SGE Assay**

| Computational Tool Prediction | Control Obs. | Case Obs. | Odds Ratio[a] | 95% C.I. | Proportion Pathogenic[b] |
|---|---|---|---|---|---|
| AGVGD | | | | | |
| Pathogenic | 8 | 2 | 3.73 | 0.77-18.1 | 0.55 |
| Indeterminate | 43 | 8 | 1.79 | 0.83-3.84 | 0.32 |
| Benign | 188 | 17 | 1.09 | 0.66-1.80 | 0.00 |
| AlphaMissense | | | | | |
| Pathogenic | 17 | 2 | 1.69 | 0.38-7.46 | 0.07 |
| Indeterminate | 115 | 15 | 1.42 | 0.82-2.45 | 0.12 |
| Benign | 107 | 10 | 1.13 | 0.59-2.17 | 0.00 |
| BayesDel | | | | | |
| Pathogenic | 77 | 13 | 1.89 | 1.04-3.44 | 0.26 |
| Indeterminate | 155 | 13 | 0.98 | 0.55-1.74 | 0.00 |
| Benign | 7 | 1 | 1.97 | 0.23-16.5 | 0.16 |
| BayesDel VCEP | | | | | |
| Pathogenic | 28 | 5 | 1.90 | 0.72-4.99 | 0.29 |
| Indeterminate | 45 | 7 | 1.79 | 0.80-4.03 | 0.21 |
| Benign | 166 | 15 | 1.07 | 0.62-1.82 | 0.00 |
| MutPred2 | | | | | |
| Pathogenic | 7 | 0 | NA | NA | NA |
| Indeterminate | 79 | 11 | 1.43 | 0.75-2.71 | 0.15 |
| Benign | 153 | 16 | 1.28 | 0.76-2.15 | 0.02 |
| REVEL | | | | | |
| Pathogenic | 85 | 11 | 1.45 | 0.77-2.74 | 0.11 |
| Indeterminate | 154 | 16 | 1.23 | 0.73-2.07 | 0.02 |
| Benign | 0 | 0 | NA | NA | NA |
| VEST4 | | | | | |
| Pathogenic | 15 | 4 | 3.94[c] | 1.28-12.2 | 0.61 |
| Indeterminate | 139 | 19 | 1.40 | 0.86-2.28 | 0.14 |
| Benign | 85 | 4 | 0.66[c] | 0.24-1.82 | 0.00 |

a. Adjusted for race/ethnicity.
b. The OR threshold used for this calculation is that of moderate risk variants (OR = 2 for breast cancer and OR = 2.9 for ovarian cancer) adjusted for observational inflation in our data to OR = 2.31 and OR = 3.83 respectively.
c. Statistically significant with a p-value less than 0.05.

**Table S7: Results of Multivariate Logistic Regression**

| Analysis | Assay Coefficient[a] | Assay P-Value | Computational Tool Coefficient[a,b] | Computational Tool P-Value | Meets ACMG Expectations?[c] |
|---|---|---|---|---|---|
| SGE Functional Assay Only | 2.48 | 2.0*10^-16[d] | NA | NA | NA |
| Assay + AGVGD | 2.02 | 5.2*^-14[d] | 0.96 | 0.0033[d] | Yes |
| Assay + AlphaMissense | 1.72 | 1.0*10^-7[d] | 1.24 | 0.0002[d] | Yes |
| Assay + BayesDel | 2.18 | 2.0*10^-16[d] | 0.02 | 0.9669 | No |
| Assay + BayesDel – VCEP | 2.22 | 1.3*10^-13[d] | 0.54 | 0.0451 | No |
| Assay + MutPred2 | 2.28 | 4.0*10^-13[d] | 0.42 | 0.2677 | No |
| Assay + REVEL | 2.36 | 2.0*10^-16[d] | 0.31 | 0.1125 | No |
| Assay + VEST4 | 1.67 | 3.03*10^-8[d] | 1.70 | 7.3*10^-7[d] | No[e] |

a. Coefficients are reported as log odds with base e.
b. This coefficient was calculated in the direction of benign computational tool prediction to pathogenic computational tool prediction. If done from pathogenic prediction to benign prediction, the magnitude of the coefficient would be different, but the p-value would be the same.
c. To meet this criterion, the computational tool must stratify the data in a way that usefully adds information on top of both functionally wild-type and functionally loss-of-function results.
d. Results are statistically significant (2-sided p-value < 0.007, an appropriate threshold to account for multiple testing).
e. Despite the low p-value, pathogenic predictions from VEST4 combined with loss-of-function results from the SGE functional assay failed to meet the expected proportions pathogenic (Table 3).

**Table S8. Comparison of ACMG Points Generated by Combining Loss of Function Functional Results with Computational Tool Predictions at the +1 Supporting Pathogenic and +2 Moderate Pathogenic Thresholds.**

| | ≥+1 Pathogenic Predictions[a] | | ≥+2 Pathogenic Predictions[a] | |
|---|---|---|---|---|
| Computational Prediction | Proportion Pathogenic | ACMG Points | Proportion Pathogenic | ACMG Points |
| AlphaMissense | | | | |
| Pathogenic | 0.85 | +5 | 0.86 | +5 |
| Indeterminate | 0.17 | 0 | 0.87 | +5 |
| Benign | NA | NA | NA | NA |
| BayesDel | | | | |
| Pathogenic | 0.75 | +4 | 0.78 | +4 |
| Indeterminate | 0.96 | ≥+5 | 0.55 | +3 |
| Benign | NA | NA | NA | NA |
| MutPred2 | | | | |
| Pathogenic | 0.79 | +4 | 0.87 | +5 |
| Indeterminate | 0.75 | +4 | 0.73 | +4 |
| Benign | 0.29 | +1 | 0.29 | +1 |
| REVEL | | | | |
| Pathogenic | 0.74 | +4 | 0.71 | +4 |
| Indeterminate | 0.99 | ≥+5 | 0.83 | +5 |
| Benign | NA | NA | NA | NA |
| VEST4 | | | | |
| Pathogenic | 0.81 | +4 | 0.88 | +5 |
| Indeterminate | 0.53 | +3 | 0.61 | +3 |
| Benign | NA | NA | NA | NA |

a. These thresholds are those established in Pejaver et al. that are sufficient for each tool for either at least +1 or at least +2 ACMG point(s). In the ≥+2 analysis, variants that met the threshold for +1 ACMG points but did not meet the threshold for ≥+2 ACMG points were counted as “Indeterminate”.
