## Supplemental Methods for "When Two Plus Four Does Not Equal Six: Combining Computational and Functional Evidence to Classify BRCA1 Key Domain Missense Substitutions"

### Estimating the proportion of pathogenic variants seen in sets of cases and controls based on counts of individuals who carry variants


#### Abstract

- This Supplement finds estimators for the proportion of variants seen in a case-control study that are pathogenic.
- As an initial step, we derive maximum likelihood estimates of the proportion of variants that exist in the population that are pathogenic, and the rate at which variants are observed in an individual.
- The observations are assumed to be the number of cases and the number of controls that carry at least one variant.
- We assume that the probability of disease to carriers and non-carriers of pathogenic variants are known. The former can be derived if the latter and either of the relative risk or odds ratio are known.
- It is not necessary to make the assumption that variation rates are small.
- The estimate of the proportion of pathogenic variants observed in the sample assumes that the probability that any particular variant is seen in an individual is small.


### 1 Method

#### 1.1 Model

Assume that genetic variants occur as a Poisson process along the gene or domain in question with rate $\lambda$. Thus, if $V$ is the number of variants that a randomly chosen individual has in the region

$$V \sim \text{Poisson}(\lambda)$$

so

$$P(V = v) = \frac{\lambda^v e^{-\lambda}}{v!}$$

and, in particular,

$$P(V = 0) = e^{-\lambda}. \tag{1}$$

Assume also that the proportion of variants that are pathogenic is $p$, which is, initially, the quantity we wish to estimate. If $W$ is the number of pathogenic variants that an individual carries, then $W$ is a thinned Poisson random variable, which is itself Poisson, thus the distribution of $W$ is

$$W \sim \text{Poisson}(p\lambda)$$

so

$$P(W = w) = \frac{(\lambda p)^w e^{-p\lambda}}{w!}$$

and, in particular,

$$P(W = 0) = e^{-p\lambda}. \quad (2)$$

Let $\delta_0$ be the frequency of the disease among individuals who do not carry a pathogenic variant, and let $\delta_1$ be the frequency among pathogenic variant carriers. Typically, the relationship between these two probabilities is expressed as either the *relative risk*

$$\rho = \frac{P(\text{Disease}|\text{Exposure})}{P(\text{Disease}|\text{No exposure})} = \frac{\delta_1}{\delta_0} \quad (3)$$

or the *odds ratio*

$$\varphi = \frac{\text{Odds of disease for exposed individual}}{\text{Odds of disease for unexposed individual}} = \frac{\delta_1/(1-\delta_1)}{\delta_0/(1-\delta_0)}. \quad (4)$$

Thus, the probability that a carrier of a pathogenic variant has the disease can be calculated as either of

$$\delta_1 = \rho\delta_0 \quad (5)$$

or

$$\delta_1 = \frac{\phi\delta_0}{1 + \delta_0(\phi - 1)} \quad (6)$$

depending on which may be available. Note that the odds ratio can be estimated from a contingency table for a case-control study while the relative risk requires random population sampling, or knowledge of population carrier probabilities.

Note that $p$ is the proportion of all variants that are pathogenic, regardless of whether they've been observed or not. However, in case-control data sets, cases and, hence, pathogenic variants will typically be over represented in comparison with the population. Thus, we use $q$ to denote the proportion of variants observed in a case-control study that are pathogenic. Typically, we expect $q > p$.

#### 1.2 Derived parameters

For the above model, we can derive the following parameter relationships. The full justifications are given in appendix A.

Appendix A.1 shows that the population frequency of the disease is

$$\delta = \delta_0 e^{-p\lambda} + \delta_1(1 - e^{-p\lambda}), \tag{7}$$

the probability that a control carries a variant is

$$\alpha = 1 - \frac{(1-\delta_0)e^{-\lambda}}{1-\delta}, \tag{8}$$

and the probability that a case carries a variant is

$$\beta = 1 - \frac{\delta_0 e^{-\lambda}}{\delta}. \tag{9}$$

In appendix A.2 we invert the relationship between $(\alpha,\beta)$ and $(\lambda,p)$ to get

$$\lambda = \log \frac{(1-\beta)(1-\delta_0) + (1-\alpha)\delta_0}{(1-\alpha)(1-\beta)} \tag{10}$$

and

$$p = \frac{1}{\lambda} \log \frac{(\delta_1 - \delta_0)[(1-\beta)(1-\delta_0) + (1-\alpha)\delta_0]}{(1-\beta)\delta_1(1-\delta_0) - (1-\alpha)\delta_0(1-\delta_1)}. \tag{11}$$

Then in appendix A.3 we derive an expression for a good approximation for the proportion of variants observed in a set of $n$ controls and $m$ cases that are pathogenic as

$$q \approx \frac{p[n\delta(1-\delta_1) + m(1-\delta)\delta_1]}{p[n\delta(1-\delta_1) + m(1-\delta)\delta_1] + (1-p)\delta(1-\delta)(n+m)} \tag{12}$$

#### 1.3 Maximum likelihood estimation

In an experiment, we sample at random a set of $n$ controls and $m$ cases. Let $X$ be the count of controls that carry at least one variant, and let $Y$ be the count of cases that do. Thus,

$$X \sim \text{Binomial}(n,\alpha)$$
$$Y \sim \text{Binomial}(m, \beta). \tag{13}$$

From standard Binomial estimation, we know that the maximum likelihood estimates, or *MLEs*, of $\alpha$ and $\beta$ are, respectively, $\hat{\alpha} = \frac{x}{n}$ and $\hat{\beta} = \frac{y}{m}$. Since there is a one-to-one relationship between values of $(\alpha,\beta)$ and $(p,\lambda)$, the standard likelihood machinery tells us that the MLE of a function is the function of the MLE, and we can plug these unconstrained MLEs $\hat{\alpha}$ and $\hat{\beta}$ into equations (10) and (11) get the MLEs of $\lambda$ and $p$ respectively. We then plug these into (7) and (12) to estimate $\delta$ and hence $q$.

It is possible that the MLE of $p$ occurs at a boundary of the allowable (0,1) values for a probability. If the likelihood is maximized at 0, $\hat{p}$ will be less that 0 in which case it is set to 0. If the likelihood is maximized at 1, $\hat{p}$ will be greater that 1 in which case it is set to 1.

To summarize, with the values of $\delta_0$ and $\delta_1$ assumed known, the values of $n$ and $m$ known by design, and $x$ and $y$ observed, the complete MLE process is so set

$$\hat{\alpha} = \frac{x}{n}$$

$$\hat{\beta} = \frac{y}{m}$$

$$\hat{\lambda} = \log \frac{(1-\hat{\beta})(1-\delta_0) + (1-\hat{\alpha})\delta_0}{(1-\hat{\alpha})(1-\hat{\beta})}$$

$$\hat{p} = \frac{1}{\hat{\lambda}} \log \frac{(\delta_1 - \delta_0)[(1-\hat{\beta})(1-\delta_0) + (1-\hat{\alpha})\delta_0]}{(1-\hat{\beta})\delta_1(1-\delta_0) - (1-\hat{\alpha})\delta_0(1-\delta_1)}$$

$$\text{if } \hat{p} < 0 \text{ set } \hat{p} = 0, \text{ if } \hat{p} > 1 \text{ set } \hat{p} = 1 \tag{14}$$

$$\hat{\delta} = \delta_0 e^{-\hat{p}\hat{\lambda}} + \delta_1(1 - e^{-\hat{p}\hat{\lambda}})$$

$$\hat{q} = \frac{\hat{p}[n\hat{\delta}(1-\delta_1) + m(1-\hat{\delta})\delta_1]}{\hat{p}[n\hat{\delta}(1-\delta_1) + m(1-\hat{\delta})\delta_1] + (1-\hat{p})\hat{\delta}(1-\hat{\delta})(n+m)}. \tag{15}$$

#### 1.4 Bootstrap variance estimates and confidence intervals

Rather than deriving algebraic expressions for the variances of $\hat{p}$ and $\hat{q}$, we can use the general bootstrap resampling approach. In principle, this is done by resampling some number $b$ of new sets of controls and cases uniformly at random with replacement independently from the respective observed sets, and obtaining a series of bootstrap estimates $\{\hat{p}_j\}$ and $\{\hat{q}_j\}$ for $j = 1...b$ from the resampled control and case variant carrier counts using (14) and (15). The bootstrap variances are then the sample variances of these resampled estimates. We can also get bootstrap confidence intervals from the sample quantiles.

Note that, in this situation, resampling the complete data sets and calculating the counts of variant carriers is exactly equivalent to resampling the counts of variant carriers directly using

$$X_j \sim \text{Binomial}(n, \frac{X}{n})$$

$$Y_j \sim \text{Binomial}(m, \frac{Y}{m}) \qquad \text{for } j = 1,...b \tag{16}$$

which both simplifies the process and allows us to bootstrap using only the sufficient statistics without needing access to the complete data sets.

In the example analysis seen below, for some of the sub samples of the data, the MLEs occur at or near the boundaries, that is, we see $\hat{p}$ or $\hat{q}$ at 0 or 1, in which case the usual approximate large sample confidence intervals give values outside (0,1). For that reason, we use the quantile approach to obtain bootstrap confidence intervals.

##### 1.5 Implementation

The above MLE and bootstrap processes have been implemented in the `R` statistical environment (R Core Team 2015) as described in appendix B. The code is available in a file called `numpath.R` and can be imported into an `R` session using the `source()` command

```
> source("numpath.R")
```

making all the functions available within that environment. The source file also contains the `R` code needed to perform all the analyses and shown in section 2.

#### 2 Results

##### 2.1 Example using BRCA1 data

Here is some `R` code to find the the MLEs for each of the 5 categories Bayes-1 to Bayes-5 and the combined data. The code uses functions defined in appendix B.

First we define the observed data.

```
n = 169933 m =
126455 x =
c(8,173,64,70,12) x
= c(x,sum(x)) y =
c(4,124,74,288,261)
y = c(y,sum(y))
```

We then set the risk to unexposed individuals to $\delta_0 = 0.12$ and obtain the risk to exposed individuals from the odds ratio which is set to 5.

```
d0 = 0.12
oddsrat = 5
d1 = probfromoddsratio(d0,oddsrat)
```

Then, run the analysis to get the proportions of all and observed variants that are pathogenic for each group.

```
propall = propallpath(x,n,y,m,d0,d1)
propobs = propobspath(x,n,y,m,d0,d1)
```

Then to do the bootstrapping to get standard errors and 95% confidence intervals from the resampled quantiles, we first set the quantiles that we want and the number of bootstrap sample to do.

```
lolim = 0.025
hilim = 0.975
nboots = 10000
```

Now resample estimates of the proportion of all variants that are pathogenic.

```
seall =
rep(0,length(x)) loall
= rep(0,length(x))
hiall =
rep(0,length(x)) for (i
in 1:length(x))
{boot = bootall(x[i],n,y[i],m,d0,d1,nboots) seall[i]
       = sqrt(var(boot)) loall[i] =
       quantile(boot,lolim) hiall[i] =
       quantile(boot,hilim)
}
```

Then run the same bootstrapping process for the proportion of observed variants that are pathogenic.

```
seobs = rep(0,length(x))
loobs = rep(0,length(x))
hiobs = rep(0,length(x))

for (i in 1:length(x))
{
       boot =
bootobs(x[i],n,y[i],m,d0,d1,nboots)
seobs[i] = sqrt(var(boot)) loobs[i] =
quantile(boot,lolim) hiobs[i] =
quantile(boot,hilim) }
```

We then combine the input data and results into the same matrix.

```
res = cbind(x,n,y,m, propall,seall,loall,hiall,
propobs,seobs,loobs,hiobs) colnames(res) = c("x","n","y","m",
              "P all","SE all","Lo all","Hi all",
              "P obs","SE obs","Lo obs","Hi obs")
rownames(res) =
c("Bayes1","Bayes2","Bayes3","Bayes4","Bayes5","Combined")
```

Here are the group estimates and 95% confidence intervals. The proportions are all shown as percentages. We're going to use the `xtable` package to present this.

```
showres = res[,-c(1:4)]
showres = 100*showres
require(xtable)
xtable(showres,caption="Table of results")
```

Note that it makes sense that the proportion of pathogenic variants in the sample is greater than the proportion of pathogenic variants that exist because the sample is about 42% cases, whereas, we have assumed the population frequency of cases is only about 12%. That is, we have selected strongly for cases and, consequently, pathogenic variants.

| | P all | SE all | Lo all | Hi all | P obs | SE obs | Lo obs | Hi obs |
|---|---|---|---|---|---|---|---|---|
| Bayes1 | 0.00 | 12.01 | 0.00 | 39.93 | 0.00 | 15.71 | 0.00 | 54.87 |
| Bayes2 | 0.00 | 2.19 | 0.00 | 7.59 | 0.00 | 3.74 | 0.00 | 12.87 |
| Bayes3 | 19.22 | 8.75 | 4.05 | 38.18 | 30.32 | 11.82 | 6.91 | 53.54 |
| Bayes4 | 100.00 | 3.42 | 87.33 | 100.00 | 100.00 | 1.92 | 92.88 | 100.00 |
| Bayes5 | 100.00 | 0.00 | 100.00 | 100.00 | 100.00 | 0.00 | 100.00 | 100.00 |
| Combined | 62.00 | 4.84 | 52.82 | 71.76 | 74.85 | 3.92 | 67.25 | 82.53 |

Table 1: Table of results

We also have set of plots for each data group showing the profile log likelihood functions for $p$, with $\lambda$ set at its maximizing value $\lambda\hat{}$. The red lines at the values of $\hat{}p$ confirms that this estimator really does maximize the likelihood.

```
par(mfrow=c(3,2))
for (i in
1:nrow(res))
{
       llplot(res[i,1],res[i,2],res[i,3],res[i,4]
,d0,d1) title(rownames(res)[i]) }
```

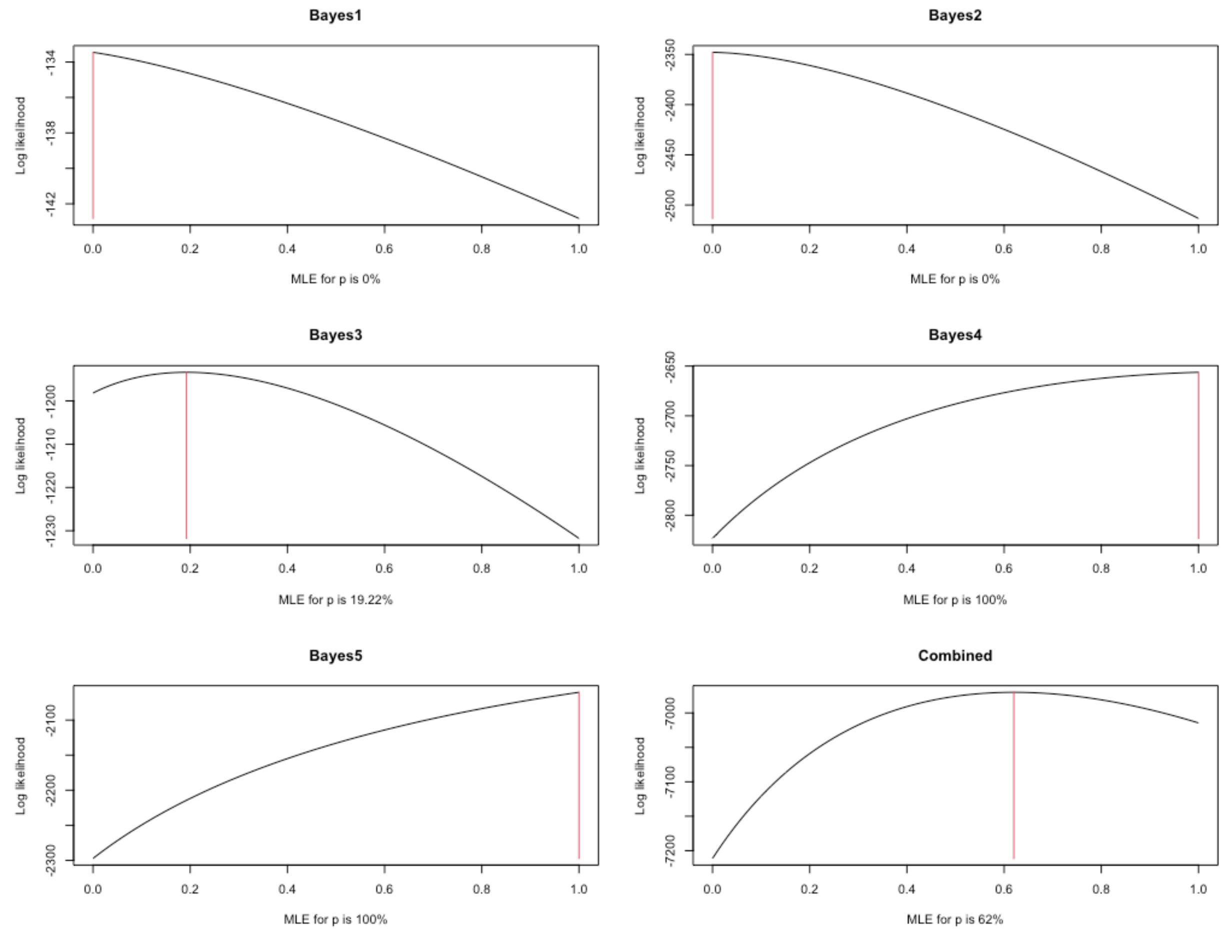


#### 2.2 Comparison with linear interpolation of odds ratio

We now compare the above method against a previously used rule of thumb method that assumes the proportion of pathogenic observed variants is 0 when the observed odds ratio,

$$\frac{Y/(m-Y)}{X/(n-X)}, \tag{17}$$

is 1, is 1 when the observed odds ratio hits the level expected for truncating variants, and interpolates linearly between these two extremes.

The following plot compares these two methods for the BRCA1 data above where we set the truncating odds ratio to 5. The odds ratio interpolation is shown as a black line. We then chose a broad set of values for $X$ and $Y$, the number of observed variant carriers in the controls and cases respectively. and computed the observed odds ratio and the proportion pathogenic estimates for these values. Interpolating lines between these points are shown in red.

We can see that the linear interpolation generally underestimates the proportion pathogenic, however, note that even though we used a broad range of $X$ and $Y$, the points calculated lie approximately on the same curve. That they don't lie on exactly the same curve is shown by the close up in the second plot. This indicates that, although a linear interpolation of the observed odds ratio is not a good estimator of the proportion of pathogenic observed variant, a very good approximation based on the odds ratio could be possible. In fact, the final plot shows that even linear interpolation using the log odds ratio is somewhat better than using the odds ratio itself.

We're going to hide the `R` code for this section for the sake of clarity.

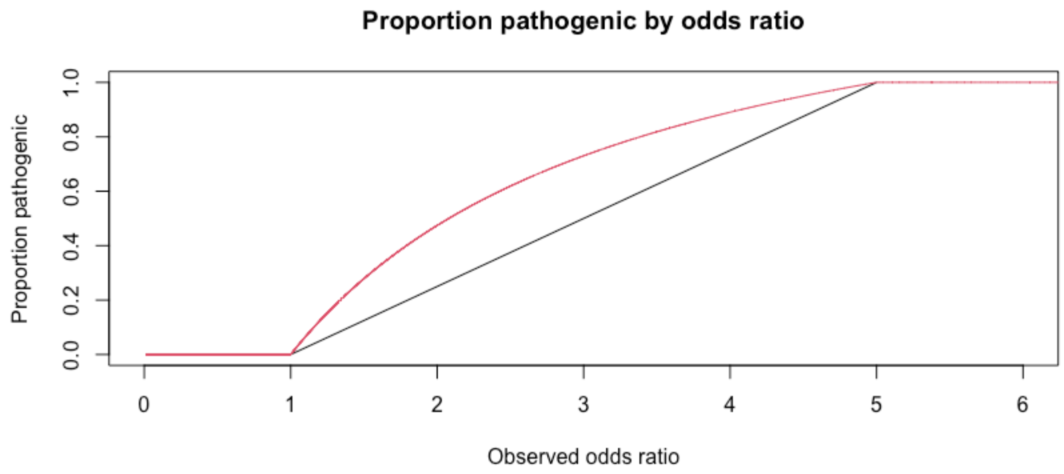


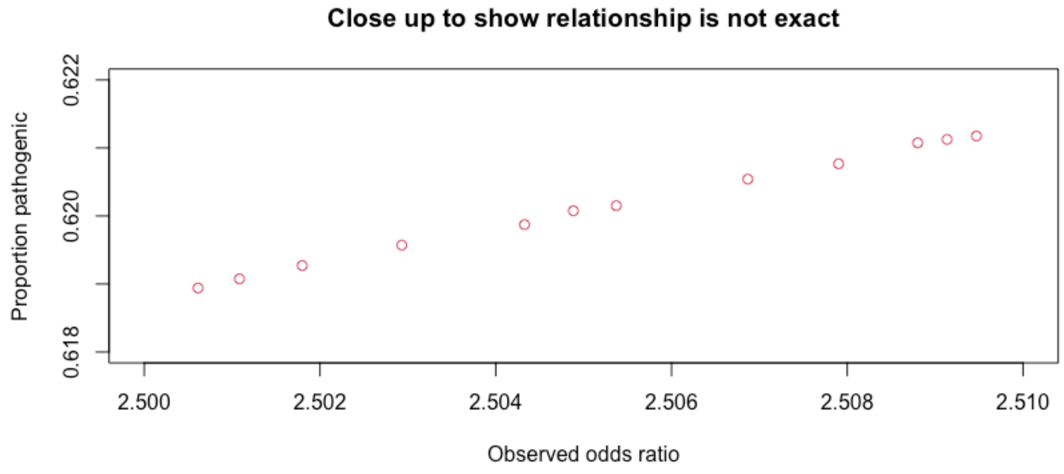

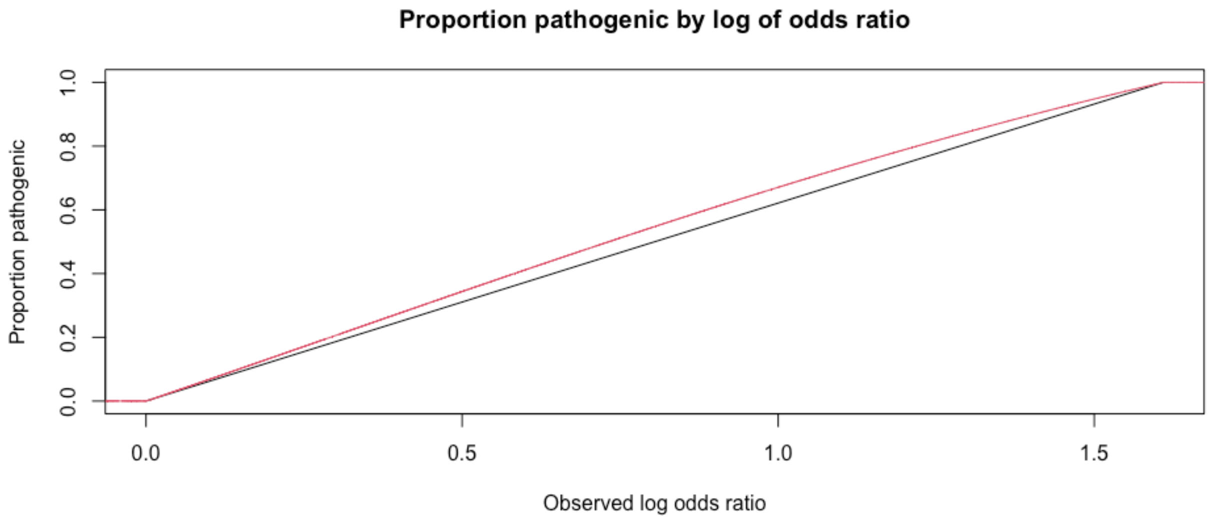

Proportion pathogenic by log of odds ratio
Proportion pathogenic
0.0
0.2
0.4
0.6
0.8
1.0
0.0
0.5
1.0
1.5
Observed log odds ratio

#### 2.3 Application to computing odds pathogenic

We consider here and application of the above method to compute estimates and bootstrap confidence intervals for the odds of pathogenicity of each subgroup with the overall total as a baseline. That is, if $p_i$ is the estimated proportion of observed pathogenic for each subgroup $i$ and $p$ proportion for the combined set, we want

$$OP_i = \frac{p_i}{(1-p_i)} \times \frac{(1-p)}{p} \qquad \forall i. \tag{18}$$

The following function will give estimates for the odds pathogenic for each group.

```
oddspathobs = function(x,n,y,m,d0,d1)
{
        p = propobspath(x,n,y,m,d0,d1) q =
        propobspath(sum(x),n,sum(y),m,d0,d
        1) p / (1-p) / q * (1-q)
}
```

To simplify things, we make a function to take a single bootstrap sample for a vector of observations.

```
oneboot = function(x,n)
{rbinom(length(x),n,x/n) }

# Check for
correctness. x =
c(1,2,3,4,100,1000)
n = 10000
b = matrix(nrow=length(x),ncol=1000)
for (i in 1:ncol(b))
        b[,i] =
        oneboot(x,n)
apply(b,1,mean)

## [1]  1.065   2.002   2.976  4.018  99.703  998.787
```

We can then combine these into a function that returns estimates with bootstrap standard errors and confidence intervals.

```
bootoddspathobs =
function(x,n,y,m,d0,d1,nb=1000,prob=0.95) {
        Group = 1:length(x)
        PropPath = propobspath(x,n,y,m,d0,d1)
```

```
        OddsPath = oddspathobs(x,n,y,m,d0,d1)
        #res =
        data.frame(Group,PropPath,OddsPath)
        res = data.frame(PropPath,OddsPath)

        b =
        matrix(nrow=length(x),ncol=nb)
        for (i in 1:ncol(b))
        {b[,i] = oddspathobs(oneboot(x,n),n,oneboot(y,m),m,d0,d1)
        }

#boxplot(t(b),ylim=c(0,20)); res$StdErr =
        sqrt(apply(b,1,var,na.rm=TRUE))

        a = (1-prob)/2
        res$CILow = apply(b,1,quantile,probs=a,na.rm=TRUE)
        res$CIHigh = apply(b,1,quantile,probs=1-a,na.rm=TRUE)

        res
}
```

Here is an example of the functions running on a test data set.

```
x = c(348,256,26,20)
n = 90832 y =
c(275,216,26,45)
m = 64044
d0 = 0.12
oddsrat =
2.41
d1 = probfromoddsratio(d0,oddsrat)

bootoddspathobs(x,n,y,m,d0,d1,2000,0.95)

##   PropPath OddsPath   StdErr      CILow  CIHigh
## 1 0.1294208 0.4883604 0.3056032 0.00000000
1.114435
##  2  0.2045590  0.8448028  0.4817115  0.01249195
1.979927 ## 3 0.4004522 2.1941776 NaN 0.00000000
Inf
## 4 1.0000000     Inf      NaN 10.58363997    Inf
```

To check these numbers you'd need the combined estimated observed proportion pathogenic.

```
q =
propobspath(sum(x),n,sum(y),m,d0,d1
) q

## [1] 0.2333683

p = 0.4004522
p/(1-p) / q * (1-
q) ## [1]
2.194177
```

### A Derivations of parameter relationships

Throughout this appendix we use notation and variable names defined in the main text.

#### A.1 Probabilities that controls and cases carry a variant

We first express the probability of $D$, the event that a randomly chosen individual from the population has the disease, in terms of our model parameters as

$$\begin{aligned}\delta &= P(D)\\ &= P(D|W=0)P(W=0) + P(D|W>0)P(W>0)\\ &= \delta_0 e^{-p\lambda} + \delta_1(1 - e^{-p\lambda}).\end{aligned} \tag{18}$$

We can now calculate the probability that a case and a control carry at least 1 variant. For controls

$$\begin{aligned}\alpha &= P(V > 0|\bar{D})\\ &= 1 - P(V = 0|\bar{D})\\ &= 1 - \frac{P(\bar{D}|V = 0)P(V = 0)}{P(\bar{D})}\\ &= 1 - \frac{(1 - P(D|V = 0))P(V = 0)}{1 - P(D)}\\ &= 1 - \frac{(1 - \delta_0)e^{-\lambda}}{1 - \delta},\end{aligned} \tag{19}$$

and similarly, for cases

$$\begin{aligned}\beta &= P(V > 0|D)\\ &= 1 - P(V = 0|D)\\ &= 1 - \frac{P(V = 0, D)}{P(D)}\\ &= 1 - \frac{P(D|V = 0)P(V = 0)}{P(D)}\\ &= 1 - \frac{\delta_0 e^{-\lambda}}{\delta}.\end{aligned} \tag{20}$$

#### A.2 Inverting the relationship

We now show how $p$ and $\lambda$ can be expressed in terms of $\alpha$ and $\beta$.

To find $\lambda$, using equations (19) and (20), express $e^{-\lambda}$ in terms of $\alpha$ and $\beta$ to get

$$e^{-\lambda} = \frac{(1-\alpha)(1-\delta)}{1-\delta_0}$$

$$e^{-\lambda} = \frac{(1-\beta)\delta}{\delta_0} \tag{21}$$

so

$$\frac{(1-\alpha)(1-\delta)}{1-\delta_0} = \frac{(1-\beta)\delta}{\delta_0}$$

and

$$\frac{\delta}{1-\delta} = \frac{(1-\alpha)}{(1-\beta)}\frac{\delta_0}{(1-\delta_0)}$$

giving

$$\delta = \frac{(1-\alpha)\delta_0}{(1-\beta)(1-\delta_0)+(1-\alpha)\delta_0}. \tag{22}$$

We can remove $\delta$, which depends on $p$, from equation (21) to get

or

$$(1-\beta)(1-\delta_0)+(1-\alpha)\delta_0$$

$$\lambda = \log\frac{(1-\beta)(1-\delta_0)+(1-\alpha)\delta_0}{(1-\alpha)(1-\beta)}.$$

$$e^{-\lambda} = \frac{(1-\beta)}{\delta_0}\frac{(1-\alpha)\delta_0}{(1-\beta)(1-\delta_0)+(1-\alpha)\delta_0}$$

$$e^{-\lambda} = \frac{(1-\alpha)(1-\beta)}{\phantom{x}}$$

and so

(23)

To find $p$, we note that from equation (18) we also have

$$\delta = \delta_0 e^{-p\lambda} + \delta_1(1 - e^{-p\lambda})$$

$$= \delta_0 e^{-p\lambda} + \delta_1 - \delta_1 e^{-p\lambda}$$

or

$$\delta_0 e^{-p\lambda} + \delta_1(1 - e^{-p\lambda}) = \delta_0 e^{-p\lambda} + \delta_1 - \delta_1 e^{-p\lambda}.$$

thus, $$e^{-p\lambda}(\delta_1 - \delta_0) = \delta_1 - \delta$$
$$= \delta_1 - \frac{(1-\alpha)\delta_0}{(1-\beta)(1-\delta_0) + (1-\alpha)\delta_0}$$
$$= \frac{\delta_1(1-\beta)(1-\delta_0) + \delta_1(1-\alpha)\delta_0 - (1-\alpha)\delta_0}{(1-\beta)(1-\delta_0) + (1-\alpha)\delta_0}$$
$$= \frac{(1-\beta)\delta_1(1-\delta_0) - (1-\alpha)\delta_0(1-\delta_1)}{(1-\beta)(1-\delta_0) + (1-\alpha)\delta_0},$$

and so, $$p = \frac{1}{\lambda}\log\frac{(\delta_1 - \delta_0)[(1-\beta)(1-\delta_0) + (1-\alpha)\delta_0]}{(1-\beta)\delta_1(1-\delta_0) - (1-\alpha)\delta_0(1-\delta_1)} \quad (24)$$

#### A.3 Observed variants

So far, we've considered $p$ the proportion of all variants that are pathogenic, regardless of whether they've been observed or not, however, in case-control data sets, cases and, hence, pathogenic variants will typically be over represented in comparison with the population. We now consider the proportion of variants observed in a case-control study that are pathogenic. That is, we want to find $q$ the probability that a randomly chosen variants is pathogenic, given that it has been observed in the data set under consideration,

$$q = P(\text{variant is pathogenic}|\text{variant is observed})$$
$$= \frac{P(\text{variant is pathogenic and variant is observed})}{P(\text{variant is observed})}. \quad (25)$$

First consider some random variant $i$ with population frequency $r_i$. If $\alpha_i$ and $\beta_i$ are the probabilities that a control or a case, respectively, carry variant $i$, then, with $X_i$ and $Y_i$ as the counts of variant $i$ in all controls and cases respectively

$$X_i \sim \text{Binomial}(n, \alpha_i)$$
$$Y_i \sim \text{Binomial}(m, \beta_i). \quad (26)$$

Hence, the probability that variant $i$ is observed in the data set is

$$P(i \text{ observed}) = 1 - P(X_i = 0)P(Y_i = 0)$$
$$= 1 - (1 - \alpha_i)^n(1 - \beta_i)^m. \quad (27)$$

Similarly, if $\alpha_i'$ and $\beta_i'$ are the probabilities of observation for a pathogenic variant

$$P(i \text{ pathogenic and observed}) = 1 - (1 - \alpha_i')^n(1 - \beta_i')^m \quad (28)$$

and a very good approximation for the required probability is, therefore, given by

$$P(i \text{ pathogenic}|i \text{ observed}) = \frac{1-(1-\alpha_i')^n(1-\beta_i')^m}{1-(1-\alpha_i)^n(1-\beta_i)^m} \approx \frac{n\alpha_i' + m\beta_i'}{n\alpha_i + m\beta_i}, \quad (29)$$

the approximations being the usual first order one from small probabilities.

To obtain the denominator, first consider $\alpha_i = P(\text{carries } i|\bar{D})$, the probability that a control carries variant i, for a general variant. We have to use a slightly different approach to the derivations of $\alpha$ and $\beta$ in equations (19) and (20) because if an individual does not carry variant $i$, they can still carry other pathogenic variants, thus, they have the population risk of the disease, not the non-carrier risk. So, if $V_i$ is the event that the individual carries VUS $i$ and $R_i$ is the event that variant $i$ is pathogenic

$$\begin{aligned}\alpha_i &= P(V_i|\bar{D}) \\ &= \frac{P(V_i, \bar{D})}{P(\bar{D})} \\ &= \frac{P(\bar{D}, R_i, V_i) + P(\bar{D}, \bar{R}_i, V_i)}{P(\bar{D})} \\ &= \frac{P(\bar{D}|R_i)P(R_i|V_i)P(V_i) + P(\bar{D}|\bar{R}_i)P(\bar{R}_i|V_i)P(V_i)}{P(\bar{D})} \\ &= \frac{r_i}{1-\delta}[p(1-\delta_1) + (1-p)(1-\delta)]. \end{aligned} \quad (30)$$

Similarly,

$$\begin{aligned}\beta_i &= P(V_i|D) \\ &= \frac{r_i}{\delta}[p\delta_1 + (1-p)\delta]. \end{aligned} \quad (31)$$

For the numerator, we see that in deriving $\alpha_i$ and $\beta_i$ in (30) and (31) we considered the case of $i$ pathogenic, ($R_i$), and $i$ benign, ($\bar{R}_i$), so for the numerator we just want to pick out the terms for $i$ pathogenic to get

$$\begin{aligned}\alpha_i' &= P(V_i, R_i|\bar{D}) \\ &= \frac{P(\bar{D}|R_i)P(R_i|V_i)P(V_i)}{P(\bar{D})} \\ &= \frac{(1-\delta_1)pr_i}{1-\delta} \end{aligned} \quad (32)$$

and

$$
\begin{aligned}
\beta_i' &= P(V_i, R_i|D) \\
&= \frac{P(D|R_i)P(R_i|V_i)P(V_i)}{P(D)} \\
&= \frac{\delta_1 p r_i}{\delta}.
\end{aligned} \tag{33}
$$

Thus,

$$
\begin{aligned}
P(i \text{ path}|i \text{ obs}) &\approx \frac{\frac{nr_i}{1-\delta}(1-\delta_1)p + \frac{mr_i}{\delta}\delta_1 p}{\frac{nr_i}{1-\delta}[(1-\delta_1)p + (1-\delta)(1-p)] + \frac{mr_i}{\delta}[\delta_1 p + \delta(1-p)]} \\
&= \frac{n\delta(1-\delta_1)p + m(1-\delta)\delta_1 p}{n\delta[(1-\delta_1)p + (1-\delta)(1-p)] + m(1-\delta)[\delta_1 p + \delta(1-p)]} \\
&= \frac{p[n\delta(1-\delta_1) + m(1-\delta)\delta_1]}{p[n\delta(1-\delta_1) + m(1-\delta)\delta_1] + (1-p)[n\delta(1-\delta) + m\delta(1-\delta)]} \\
&= \frac{p[n\delta(1-\delta_1) + m(1-\delta)\delta_1]}{p[n\delta(1-\delta_1) + m(1-\delta)\delta_1] + (1-p)\delta(1-\delta)(n+m)}.
\end{aligned} \tag{34}
$$

We see that this approximation in equation (34) does not depend on $r_i$ as it cancels from each term, so it is a general expression for $q$, that is,

$$
q = \frac{p[n\delta(1-\delta_1) + m(1-\delta)\delta_1]}{p[n\delta(1-\delta_1) + m(1-\delta)\delta_1] + (1-p)\delta(1-\delta)(n+m)}. \tag{35}
$$

#### B R functions to implement the methods

Here are functions to implement the methods we derived above written in the R statistical environment.

First, here are functions to obtain the frequency of disease in pathogenic variant carriers from the baseline risk and either relative risk or odds ratio.

```
probfromrelativerisk = function(delta0, rho)
{rho * delta0
}

probfromoddsratio = function(delta0, phi)
{
       phi * delta0 / ( 1 + delta0 * (phi-1))
}
```

Then some functions to express $\alpha$ and $\beta$ in terms of $p$ and $\lambda$.

```
delta = function(p,l,d0,d1)
{d0 * exp(-p*l) + d1*(1-exp(-p*l))
}

alpha = function(p,l,d0,d1)
{
       1 - (1-d0)*exp(-l) / (1-delta(p,l,d0,d1))
}

beta = function(p,l,d0,d1)
{
       1 - d0*exp(-l) /
delta(p,l,d0,d1) }
```

Now, the functions to invert the relationship giving $\lambda$ and $p$ in terms of $\alpha$ and $\beta$.

```
lambda = function(a,b,d0)
{log( ((1-b)*(1-d0) + (1-a)*d0) / (1-a)/(1-b))
}

pp = function(a,b,d0,d1)
{
```

```
p = (d1-d0) * ((1-b)*(1-d0) + (1-a)*d0) p = p / (
(1-b)*d1*(1-d0) - (1-a)*d0*(1-d1) ) log(p) /
lambda(a,b,d0)
}
```

Here is the function that finds the MLE for *p*, the proportion of extant pathogenic variants, truncating between 0 and 1 when necessary.

```
propallpath = function(x,n,y,m,d0,d1)
{
       p = pp(x/n,y/m,d0,d1)
       p[p<0] =
       0 p[p>1]
       = 1 p
}
```

Here is a function that estimates the proportion of pathogenic variants observed in the sample.

```
propobspath = function(x,n,y,m,d0,d1)
{
       p =
       propallpath(x,n,y,m,d0,d1
       ) l = lambda(x/n,y/m,d0)
       d = delta(p,l,d0,d1)

       ai = (p * (1-d1) + (1-p) * (1-d)) / (1-d)
       bi = (p * d1 + (1-p)*d) / d
       pobs = n * ai + m * bi
       aip = p * (1-d1) /(1-d)
       bip = p * d1 / d
       ppathobs = n*aip + m * bip
       ppathobs/pobs
}
```

The following functions give bootstrap samples from which we can obtain variance estimates and confidence intervals.

```
bootall = function(x,n,y,m,d0,d1,b=1000)
{
       xjs = rbinom(b,n,x/n) yjs
       = rbinom(b,m,y/m)
       propallpath(xjs,n,yjs,m,d
       0,d1)
```

```
}

bootobs = function(x,n,y,m,d0,d1,b=1000)
{
       xjs =
       rbinom(b,n,x/n) yjs
       = rbinom(b,m,y/m)
       propobspath(xjs,n,yjs,m,d0
,d1) }
```

Here is a function to calculate the log likelihood for the data in terms of $p$ and $\lambda$.

```
loglike = function(x,n,y,m,p,l,d0,d1)
{
       a =
       alpha(p,l,d0,d1)
       b =
       beta(p,l,d0,d1)
   x * log(a) + (n-x)*log(1-a) + y * log(b) + (m-y) * log(1-b)
}
```

And a function to plot the log of the profile likelihood for $p$ with $\lambda$ set to its MLE $\hat{\lambda}$. Vertical lines indicate where $\hat{p}$ is.

```
llplot = function(x,n,y,m,d0,d1)
{
       phat =
       propallpath(x,n,y,m,d0,d1)
       allp = (0:100)/100 lhat =
       lambda(x/n,y/m,d0)
       ll = loglike(x,n,y,m,allp,lhat,d0,d1)
       xl = paste("MLE for p is
",round(100*phat,2),"%",sep="")
plot(allp,ll,type="l",xlab=xl,ylab="Log likelihood")
lines(c(phat,phat),range(ll),col=2) }
```
